# ClinVec: Unified Embeddings of Clinical Codes Enable Knowledge-Grounded AI in Medicine

**DOI:** 10.1101/2024.12.03.24318322

**Authors:** Ruth Johnson, Uri Gottlieb, Galit Shaham, Lihi Eisen, Jacob Waxman, Stav Devons-Sberro, Curtis R. Ginder, Peter Hong, Raheel Sayeed, Xiaorui Su, Ben Y. Reis, Ran D. Balicer, Noa Dagan, Marinka Zitnik

## Abstract

Integrating structured clinical knowledge into artificial intelligence (AI) models remains a major challenge. Medical codes primarily reflect administrative workflows rather than clinical reason ing, limiting AI models’ ability to capture true clinical relationships and undermining their generalizability. To address this, we introduce *ClinGraph*, a clinical knowledge graph that integrates eight EHR-based vocabularies, and *ClinVec*, a set of 153,166 clinical code embeddings derived from *ClinGraph* using a graph transformer neural network. *ClinVec* provides a machine-readable representation of clinical knowledge that captures semantic relationships among diagnoses, medications, laboratory tests, and procedures. Panels of clinicians from multiple institutions evaluated the embeddings across 96 diseases and more than 3,000 clinical codes, confirming their alignment with expert knowledge. In a retrospective analysis of 4.57 million patients from Clalit Health Services, we show that *ClinVec* supports phenotype risk scoring and stratifies individuals by survival outcomes. We further demonstrate that injecting *ClinVec* into large language models improves performance on medical question answering, including for region-specific clinical scenarios. *ClinVec* enables structured clinical knowledge to be injected into predictive and generative AI models, bridging the gap between EHR codes and clinical reasoning.

## Introduction

Medicine builds on centuries of accumulated knowledge and the goal of individualized patient care through clinical reasoning and evidence-based practice^1^. Over time, medical vocabularies and ontologies have been developed to represent clinical information, fostering interoperability and data exchange across healthcare systems^2–4^. However, these standardized coding systems re main fragmented, with each vocabulary designed for specific use cases. This fragmentation limits their integration into artificial intelligence (AI) models and introduces a gap between clinical knowledge and its computational representation in precision medicine.

The widespread adoption of electronic health records (EHRs) and data standardization initiatives has accelerated the use of structured clinical data in AI models^5,6^, with more than half of healthcare foundation AI models relying on structured clinical codes, such as billing data and medication records^7^. These models implicitly assume that EHRs contain information sufficient to capture the clinical concepts relevant to patient care. In practice, this assumption often fails. Models trained on EHR data frequently learn patterns specific to a single healthcare system rather than generalizable medical relationships^8–11^. The variability of clinical codes across institutions further limits the transferability of prediction models^12–15^. Many codes are selected or recorded for operational rather than clinical reasons^16^, influenced by geographic factors^17–19^, na tional policies^20–23^, insurance requirements^24–26^, physician practices^27–29^, and other features of local healthcare delivery^30–34^. EHR-based models, as a result, often fail to reflect consistent clinical definitions or relationships among codes^35–39^.

The lack of alignment between EHR-based prediction models and clinical knowledge also introduces concerns about model bias and transparency^40^. Although large language models (LLMs) offer implicit access to clinical knowledge embedded within their training data, unvetted datasets from biological data repositories and scientific articles used to train LLMs can increase the risk of bias and mistakes in generated outputs^41–43^. LLMs can struggle to represent clinical codes^44–46^, especially rare or highly specific codes, resulting in inaccuracies, over-generalizations, or hallucinations during inference^47^. This can be concerning given that a large portion of medical codes may not be used in everyday clinical practice where, for example, fewer than 5% of SNOMED CT codes account for 95% of usage in healthcare institutions^48^, creating a mismatch between what is modeled and what is clinically relevant.

Most AI models lack mechanisms to integrate clinical knowledge, limiting their ability to align with medical best practices^49,50^. Clinical researchers have published over 3,700 evidence-based guidelines across 39 countries^51–54^, yet this structured knowledge remains largely unused in model development. Most AI models rely on statistical patterns in patient data or text corpora, but these approaches do not capture the clinical knowledge embedded in structured EHR codes. Bridging this gap requires consistent, machine-readable representations that reflect how clinical codes relate to real-world medical practice. While embeddings exist for text and images^55–60^, no comparable resource captures structured clinical knowledge for use in AI models.

To address the gap between clinical knowledge and structured EHR data, we developed a *ClinGraph*, a clinical knowledge graph spanning eight standardized medical vocabularies, and *ClinVec*, a set of 153,166 clinical code embeddings derived from *ClinGraph. ClinVec* defines a unified, machine-readable latent space that captures relationships among diagnoses, medications, laboratory tests, and procedures. This approach generates clinically meaningful representations of medical codes in a hypothesis-free manner and provides a resource for integrating structured clinical knowledge into predictive models for precision medicine. The learned embeddings reveal patterns aligned with human anatomy and disease phenotypes and were validated by panels of clinical experts across 96 diseases and more than 3,000 clinical codes. We further demonstrate the predictive utility of *ClinVec* through phenotype risk score analyses for chronic kidney disease, chronic obstructive pulmonary disease, and heart failure, using EHR data from 4.57 million patients in Clalit Health Services. Finally, we show that integrating *ClinVec* into large language models improves medical question answering across a range of clinical scenarios. *ClinGraph* and *ClinVec* provide a foundation for embedding structured clinical knowledge into AI models, supporting interpretability, cross-institutional transferability, and alignment with clinical practice.

## Results

### *ClinGraph* and *ClinVec*: Representing structured medical knowledge for AI

Many efforts have focused on structuring clinical knowledge for enhanced interoperability and data harmonization. Clinical knowledge has been traditionally structured using Common Data Models (CDMs), which standardize data by mapping diverse coding systems into a unified schema^61^. Alternatively, biomedical ontologies organize information in a hierarchical structure to facilitate semantic interoperability and reasoning across coding systems. However, both rely on fixed schemas and expert-defined relationships that limit their contextual richness and adaptability to real-world data. KGs offer a more flexible approach by integrating diverse knowledge sources without rigid schema constraints. However, most knowledge graphs do not explicitly incorporate standard clinical vocabularies and instead focus on biological processes^62–64^, therapeutics^55,65,66^, genetic phenotypes^67–70^, or a combination thereof^71–73^. KGs that incorporate clinical vocabularies are often restricted to ICD codes (diagnosis codes) or rely on medical terminologies like HPO^74^ and MONDO^75^, which are not standard in most EHRs^76–78^. Several approaches construct KGs directly from patient data^79–81^, but this process is highly sensitive to the source patient population, potentially limiting generalizability.

We develop a resource consisting of *ClinGraph*, a knowledge graph consisting of 153,166 nodes and 2,873,025 edges spanning eight clinical vocabularies (Figure 1A, 1B), and *ClinVec*, a set of high-dimensional, vector representations (embeddings) of clinical knowledge that can be integrated into AI models (Figure 1C, 1D). *ClinGraph* incorporates diagnosis codes (ICD9CM, ICD10CM, phecodes), laboratory tests (LOINC), medications (RxNorm, ATC), procedures (CPT), and other controlled clinical vocabularies (SNOMED CT, UMLS), and represents each concept in the KG as a medical code (Supplementary Table S1). Relational information is derived from multiple databases, including ontologies such as Unified Medical Language System^82^ (UMLS) and empirically constructed knowledge bases such as PheMap^83^, which derives relationships from the large-scale mining of biomedical literature (Supplementary Table S2).

**Figure 1:**
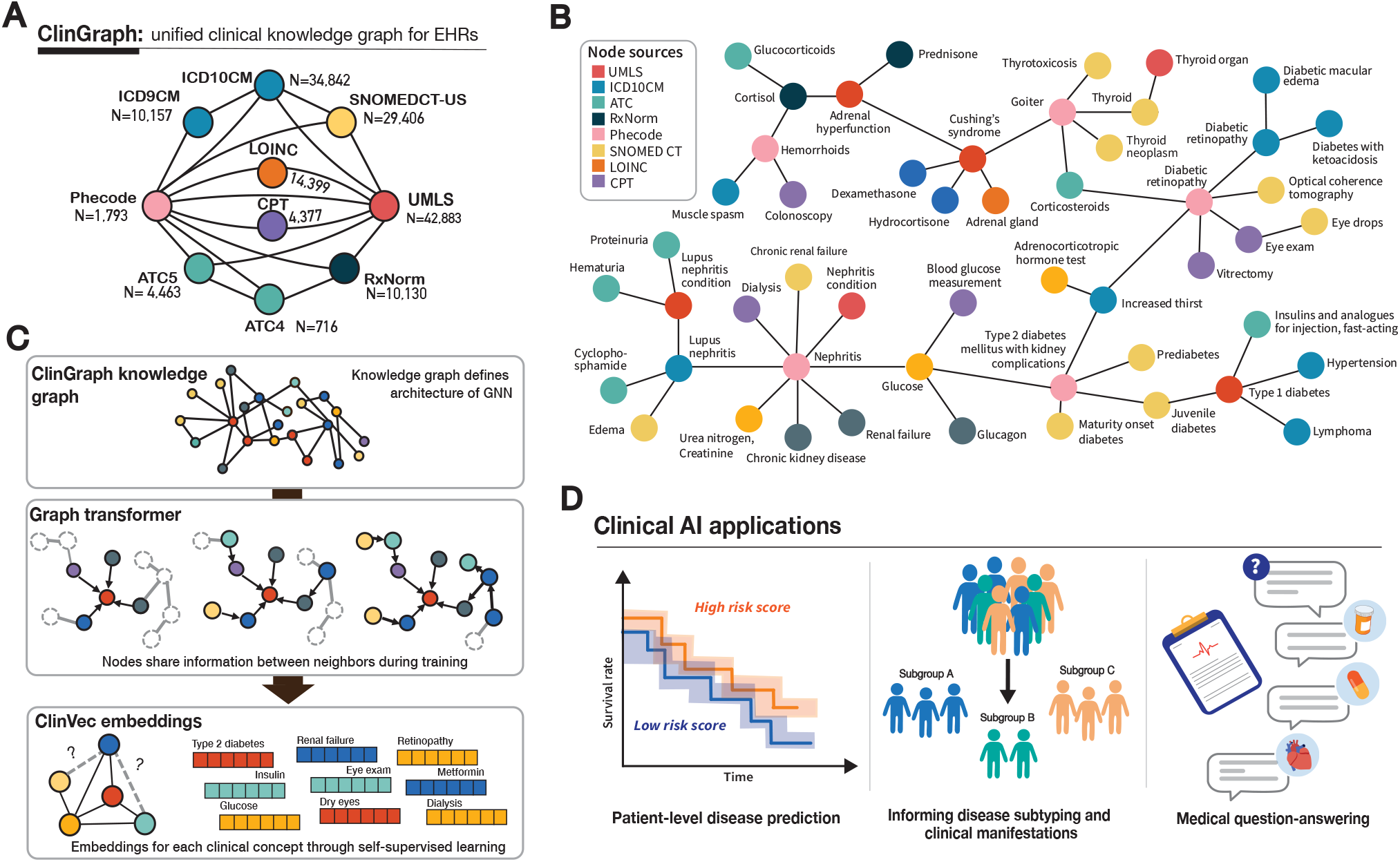
Overview of approach. **(A)** Meta-graph showing the structure of *ClinGraph* and the edge relationships between clinical vocabularies shown as node types. **(B)** An example local neighborhood of *ClinGraph*. Each clinical code is represented as a node, and the color denotes the clinical vocabulary. **(C)** Workflow for generating clinical knowledge embeddings: building the clinical knowledge graph, constructing a graph transformer model, and gener ating embeddings through self-supervised learning. **(D)** Clinical AI applications of *ClinGraph* and *ClinVec*.

A key feature of *ClinGraph* is that it represents clinical concepts using their original coding systems, rather than aggregating them into a common vocabulary. This design preserves coding-specific structures such as hierarchical relationships and clinically meaningful groupings and avoids the information loss that can occur when mapping between vocabularies. To evaluate the cover age of EHR coding systems, we compared *ClinGraph* to six existing KGs. For each KG, we mapped its node identifiers to EHR codes and calculated the proportion of coverage for common coding systems (Supplementary Table S3). *ClinGraph* is the only resource that integrates diagnoses, medications, laboratory tests, and procedures in a unified framework based on structured EHR data. Other KGs do not include nodes for procedures or laboratory tests. While *ClinGraph*, PrimeKG^76^, and PharMeBINet^84^ capture similar proportions of ICD10CM codes, *ClinGraph* spans a broader range of diagnoses. By evaluating the number of distinct ICD10CM root codes (for example, J45), we found that *ClinGraph* includes 35% more disease areas than other graphs. This likely reflects the clinical orientation of *ClinGraph*, in contrast to other resources that emphasize biological and genetic phenotypes.

While KGs represent information as discrete nodes and edges, embedding-based representations create a machine-readable form of information that encodes clinical concepts and their contextual information into continuous vectors. Transforming discrete KGs into continuous embeddings enables complex inference tasks, such as grounding AI models in clinical knowledge^85^, that are not possible using only node-and-edge graph representations. Using a heterogenous graph transformer (HGT)^86^, we learn a function that maps each node (medical code) in the KG to low-dimensional representations (embeddings). The KG determines how information is shared between nodes throughout the KG, while a multi-head attention mechanism enhances the model’s ability to up-weight and down-weight different parts of the topological space^87^. We model four distinct node types—diagnosis codes, medications, laboratory tests/procedures, and clinical concepts/vocabularies—where grouping similar vocabularies (e.g., RxNorm and ATC) enables information sharing across related nodes while preserving enough type-specific infor mation to support effective parameter learning (Methods). We train the model for edge predic tion through self-supervised learning via contrastive edge masking^88^. This approach generates embeddings for each clinical code such that codes located near one another in the knowledge graph are also positioned closely in the latent space, reflecting their semantic similarity.

To support the integration of *ClinVec* into AI models, the learned embeddings must be consistent with current clinical knowledge and reflect information used in routine healthcare. Existing reference datasets for evaluating the semantic relatedness of clinical terms are limited in size (fewer than 1,500 data points) and based on outdated coding systems that include obsolete codes. We constructed a new validation dataset consisting of 96 diseases across 10 major disease categories, such as circulatory and respiratory disorders, and over 3,000 SNOMED CT codes. A panel of expert clinicians from Israel and the United States annotated the dataset. Based on clinician input, we selected 96 phecodes representing conditions that are clinically relevant to general adult patient populations. For each phecode, we identified the 20 SNOMED CT codes with the highest cosine similarity to the phecode embedding and randomly sampled an additional 20 SNOMED CT codes as controls (Figure 2A).

**Figure 2:**
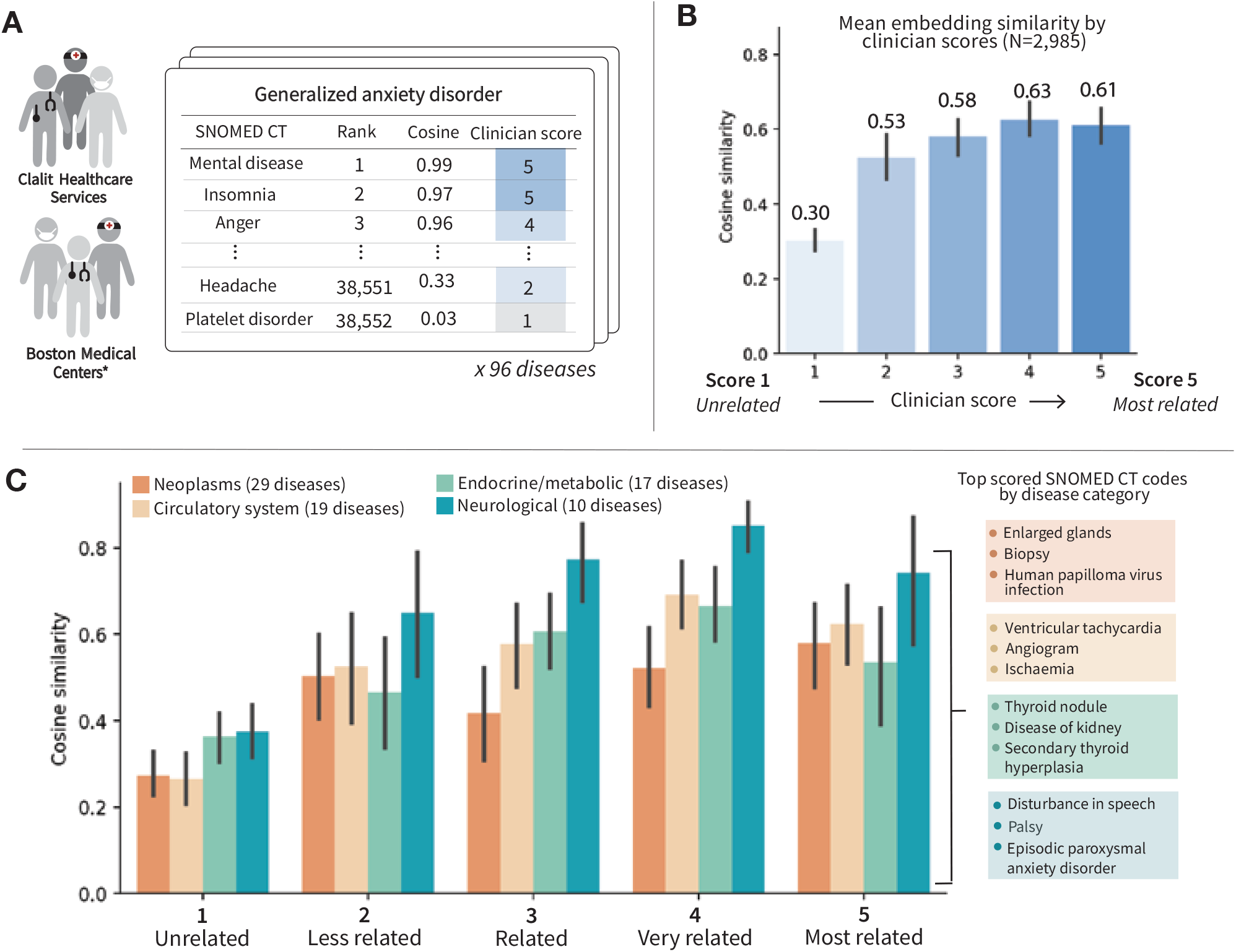
Concordance between *ClinVec* embedding similarity and clinical expert scores. **(A)** Overview of clinical expert review process. **(B)** We report the mean cosine similarity across all disease-term pairs stratified by clini cal expert scores 1-5. **(C)** We stratify cosine similarity scores across four disease categories and show examples of SNOMED CT codes that were assigned a clinical expert score of 5.

Codes already forming an edge with the disease node within the knowledge graph were excluded from the analysis. The final evaluation dataset comprises 96 disease lists, each with 40 SNOMED CT codes for a total of 3,680 data points, almost 3 times the size of existing bench marks combined.

To assess the relevance of each clinical code to a given disease, clinicians graded SNOMED CT codes on a scale from 1 (unrelated) to 5 (very relevant), evaluating both accuracy and applicability to clinical practice (Figure 2B). Clinicians graded each SNOMED CT code on a scale from 1 to 5, where 1 indicated the code was unrelated to the target disease and 5 indicated it was highly relevant and frequently observed in clinical practice. We compared these expert ratings to the cosine similarity between the corresponding *ClinVec* embeddings. Grouping similarity scores by clinician rating revealed a positive correlation, with mean cosine similarity increasing across scores 1 to 5 (0.30, 0.53, 0.58, 0.63, 0.61). When stratifying by disease category, we observed lower alignment for neoplasms compared to other categories. In this case, high embedding similarity was often assigned a low clinical relevance score by reviewers. This misalignment likely reflects the heterogeneity of neoplasms, which vary by organ system, stage, and metastatic behavior. Among disease-code pairs with low clinician scores (≤2) and high cosine similarity (>0.8), many mismatches involved anatomical proximity rather than clinical relevance. For example, top-scoring terms for “cancer of salivary glands” included thyroid conditions. Although these organs are anatomically adjacent and involved in autoimmune diseases such as Sjögren’s syndrome or Hashimoto’s thyroiditis, they are not directly related in the context of salivary gland cancer. These findings suggest that embeddings for highly specific diseases could benefit from fine-tuning on subgraphs of the KG to improve granularity and clinical alignment.

### Anatomical and medical subspecialty structure emerge in the embedding space

Clinical knowledge embeddings must capture patterns that align with human biology, disease presentation, and clinical workflows. Although *ClinGraph* shares some content with other biomedical KGs, its integration of clinical vocabularies and relationship types produces embeddings that more effectively represent clinically meaningful structure in the latent space. To evaluate this, we trained heterogeneous graph transformers on PrimeKG and a SNOMED CT-based KG using the same model architecture and training setup as for *ClinGraph*, and compared the result ing embedding spaces (Methods).

We characterize the breadth and granularity of the clinical knowledge represented in the learned embeddings. Visualizing the latent space of disease (Figure 3A, Supplementary Figure S1A-C) and drug embeddings (Figure 3B, Supplementary Figure S1D-F) generated by each KG, the embeddings derived from *ClinGraph* broadly maintain groupings of diseases by organ system, mirroring the organization of clinical care. However, embeddings produced by PrimeKG and SNOMED-CT KG do not exhibit this distinct clustering pattern. This observation is supported when comparing silhouette scores and adjusted mutual information (AMI) where embedding clusters from *ClinGraph* (silhouette: 0.54, AMI: 0.31) produce metrics at the same value or greater than clusters produced from other KGs (PrimeKG - silhouette: 0.54, AMI: 0.02; SNOMED-CT KG - silhouette: 0.15, AMI: −1e-4). Further examining the clusters of phecode embeddings from *ClinGraph*, although some phenotypes deviate from these main clusters, many deviations reflect known distinctions within clinical care. For example, most endocrine condi tions cluster together (Figure 3A: orange cluster), but some phenotypes cluster more closely with sense organ conditions (Figure 3A: green cluster). However, this small set of endocrine pheno types includes mainly eye-related conditions resulting from complications from endocrine disorders, such as diabetic retinopathy and exophthalmos.

**Figure 3:**
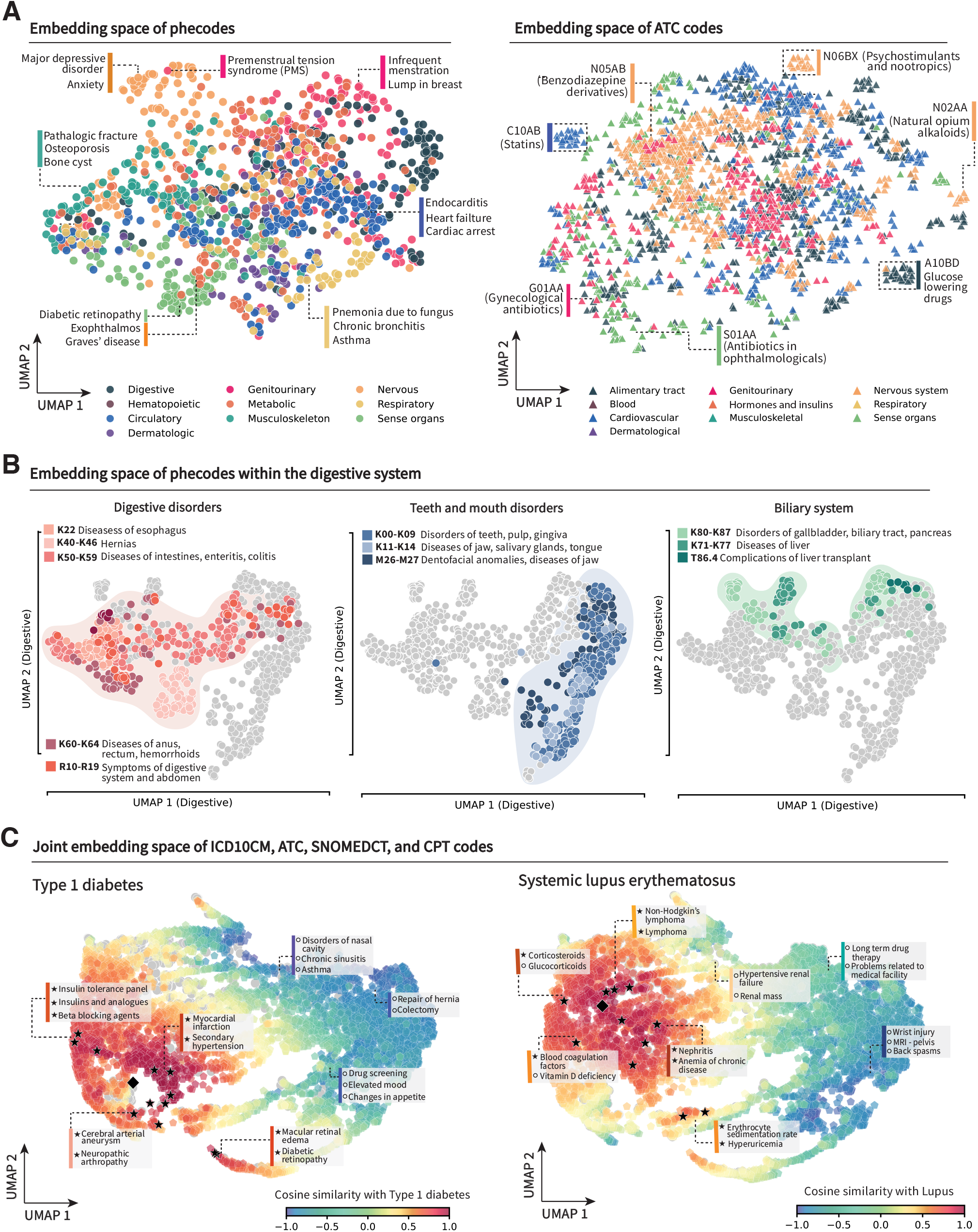
Latent *ClinVec* space captures biomedical knowledge consistent with organ system biology and anatomy. **(A)** Scatter plots showing a reduced dimension of the embedding space of clinical codes. On the left, the latent space of phecodes is shown, where each phecode is shaded by its corresponding organ system categorization. On the right, the latent space of ATC medication codes is shown, where each ATC code is shaded according to the corresponding ATC level 1. **(B)** Visualization of the embedding region of digestive system ICD10CM codes. We highlight the following phenotype subcategories: functional digestive disorders – phecodes, teeth and mouth disorders (center), and biliary system (right). **(C)** Visualization of embedding space overlaid with ICD10CM, ATC, SNOMEDCT, and CPT codes. Codes are shaded based on cosine similarity with type 1 diabetes (left) and systematic lupus erythematosus (right), and the target disease embedding is denoted with a black diamond. The black stars indicate a subset of codes related to each disease. These are presented along with other codes in their proximity but are not directly related to the disease of interest.

Extending this clustering analysis to drug categories based on the ATC-1 level, embeddings generated by both *ClinGraph* and PrimeKG show clustering by drug category although *ClinGraph* has a higher silhouette score (0.71 vs. 0.64). In contrast, embeddings from the SNOMED-CT knowledge graph do not produce distinct clusters and have an AMI score close to zero. This is likely attributable to the more extensive drug coverage in *ClinGraph* compared to SNOMED-CT KG. Again, we also find that deviations in the expected cluster labeling according to the ATC-1 classification in *ClinGraph* reflect known artifacts of the ATC coding system. For example, the embeddings for ATC-4 codes G01AA (antibiotics) and D06AX (antibiotics for topical use) are located nearby in the embedding space despite being in different ATC-1 categories. These analyses demonstrate that the effectiveness of clinical embeddings depends on the design of the underlying KG. *ClinGraph*’s integration of clinically grounded vocabularies and relationship types results in more accurate representations, outperforming existing KGs across multiple evaluation metrics.

A unified clinical vocabulary latent space must capture patterns at broad anatomical levels while also identifying discernible relationships at clinical subspecialty levels. To illustrate the granularity of *ClinVec* clinical vocabulary embeddings, we perform a separate dimensionality reduction analysis using only phecodes of the digestive system (Figure 3B). We find that conditions within the same subspecialty are significantly closer together compared with all other conditions of the digestive system, where codes describing functional digestive disorders (*p-value*=3.65×10^−45^; two-sided Mann-Whitney U test), biliary system (*p-value*=6.38×10^−44^), and conditions of mouth and teeth (*p-value*=1.05×10^−74^) are separated within the latent space. Although these codes all fall under the practice of gastroenterology, the embeddings exhibit distinct clustering patterns that reflect their unique clinical presentations and organ systems. This finding show that *ClinVec* embeddings to capture fine-grained distinctions within broader medical categories.

A key advantage of *ClinVec* embedding space is that it allows direct comparison of clinical codes across different vocabularies, bridging gaps between otherwise disconnected coding systems. We compare *ClinVec* to two state-of-the-art open-source clinical embeddings, snomed2vec^89^ and cui2vec^90^, in a set of zero-shot retrieval tasks. For a given disease, we select the top k closest codes within the embedding space and compare with these with the diseases’ symptom list as listed in the Mayo Clinic disease description. We report each model’s performance using ‘hits @ k’ which measures the proportion of diseases where a relevant symptom is ranked among the top k items within the embedding space. We find that the *ClinVec* embeddings outperform other approaches across a variety of values of *k* (Supplementary Table S4A). While hierarchical structures explain some of the clustering, the embeddings also capture meaningful patterns in cross-domain relationships, such as between diseases and their associated treatments, which are not strictly hierarchical. We perform a similar analysis for drug indications with established first-line drug treatments. For example, non-Hodgkin’s lymphoma and CHOP (cyclophosphamide, doxorubicin, vincristine, prednisone), testicular cancer and BEP (bleomycin, etoposide, cisplatin), and major depressive disorder and selective serotonin reuptake inhibitors. Limiting retrieval over codes exclusively describing ATC5-level medications, for each disease, we perform zero-shot retrieval across >2,000 drug codes and assess the codes with the highest cosine similarity with the disease embedding. We find that the *ClinVec* embeddings outperform other approaches across a variety of values of *k* (Supplementary Table S4B).

### Embedding space reveals clinically grounded diagnostic and symptom patterns

Many clinical applications require disease-level resolution, where models must distinguish between specific diagnoses and their associated symptom profiles. To evaluate how *ClinVec* captures these diagnostic patterns, we examine type 1 diabetes (T1D) and systemic lupus erythematosus (Figure 3C). We compute the cosine similarity between the T1D embedding and ICD10CM codes, CPT, LOINC, and SNOMED CT codes within the embedding space and visualize these patterns by projecting the computed cosine similarity onto the latent space. Broadly, near the embedding of T1D, we observe regions of high cosine similarity as shaded in red and orange. We find that many codes within this region are consistent with relevant T1D laboratory tests, such as ‘insulin tolerance panel’, and common complications, including cardiovascular diseases (Methods). We also identify a cluster of eye-related tests for T1D, including codes for eye exams and macular edema. This highlights the embedding space’s ability to capture nuanced patterns that extend beyond identifying codes associated with the same organ system, recapitulating patterns aligned with established disease presentations. Repeating this process for lupus, regions with high cosine similarity highlight relevant tests such as erythrocyte sedimentation rate, which is used to assess systemic inflammation and nephritis, a common complication.

*ClinVec* contains the expressivity and flexibility to describe a wide range of clinical scenarios within the embedding space. Through embedding arithmetic, embeddings can be combined mathematically to form new meanings. For example, the similarity between the vector summation of common heart disease symptoms (*e*.*g*., *shortness of breath, chest pain)* and the disease embedding of *common heart disease* yields a cosine similarity score of 0.66. These observations mirror the geometric properties of word embeddings in natural language processing^91^, effectively capturing the relationship between concepts without explicitly defining it in the original vocabulary. We refer to the aggregation of symptom embeddings in this manner as a *disease symptom embedding*—denoting that the resulting vector is a composition of the symptoms of a given disease (Figure 4A, 4B).

**Figure 4:**
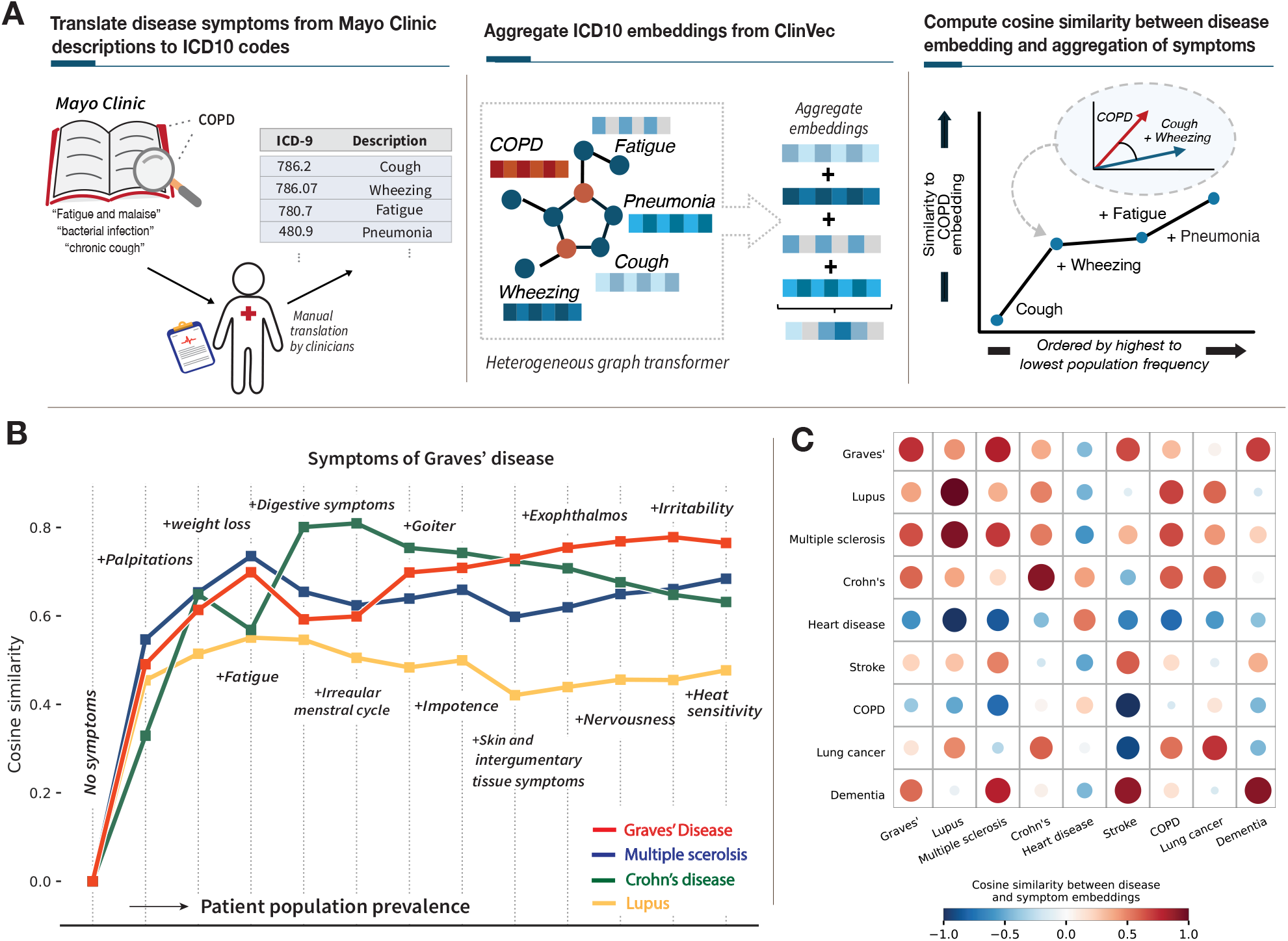
Embedding operations mirror symptomatic manifestations of clinical conditions. **(A)** Overview of the approach for constructing disease symptom embeddings. **(B)** Line plot showing the cosine similarity across four autoimmune diseases and the aggregation of Graves’ disease symptoms. Aggregated embeddings are computed by performing a vector sum across the embeddings for each symptom. **(C)** Heatmap showing cosine similarity between each disease embedding and the aggregation of symptoms. Sizes of circles correspond to the magnitude of the cosine similarity.

We assess this pattern by examining nine diseases with significant health implications across different clinical specialties. Five conditions are among the top non-communicable diseases with the highest mortality rate identified by the World Health Organization^92^ and four are among the most common autoimmune disorders in adult populations^93^. Using clinical descriptions of diseases from the Mayo Clinic^94–98^ and the manual translation of symptom lists to diagnosis codes, we aggregate all disease symptoms in the form of ICD-9 code embeddings (Supplementary Table S5). Computing the cosine similarity between the disease symptom embedding and the target disease embedding across all nine diseases, we find that all pairs have positive similarity scores (*mean-cos-similarity*: 0.77, *SD*: 0.13) (Supplementary Table S7). Moreover, conditions with overlapping symptoms also exhibit high similarity scores. For instance, the phecode embedding for multiple sclerosis also has a high similarity score with the disease composition embedding for lupus (*cos-similarity*=0.58), as both diseases share symptoms such as fatigue and muscle pain. We observe high levels of congruence between diseases affecting the same organ system, such as lung cancer (target disease) and COPD (symptom embedding), which both affect the respiratory system (*cos-similarity*=0.86).

After identifying this pattern, we evaluated whether individual symptoms exhibit consistent syntactic and semantic structure within the embedding space. To approximate the disease specificity of each symptom, we ranked symptoms by their frequency in a sample of patients from Clalit Health Services (CHS), from most to least common. For each of the nine diseases, we incrementally aggregated symptom embeddings in order of decreasing frequency. As less common symptoms were added, the combined symptom embedding progressively aligned more closely with the target disease embedding (Supplementary Figures S2, S3). To test the robustness of this trend, we perturbed key components of the embedding aggregation process. Across variations, including swapping the target disease, substituting related symptoms, and introducing randomly selected symptoms to mimic real-world variability, the symptom embedding consistently moved closer to the corresponding disease embedding in the latent space (Supplementary Figure S4).

This process can help study diseases with similar symptoms by revealing subtle differences in clinical presentations. This level of granularity can be observed when comparing various autoimmune diseases due to the presentation of overlapping symptoms. For Graves’ disease, we compute the initial cosine similarity between the disease embedding for each of the four diseases and the first symptom embedding, “palpitations”, (the Graves’ symptom found to be most prevalent in the sampled patient population), but no disease has a cosine similarity score > 0.40 (Figure 3B). Given the relatively high prevalence of this symptom in the general patient population and lack of disease specificity (*freq*=0.232), it is unsurprising that initial symptoms do not show a high correlation with any of the immune conditions. However, as we add symptoms that are relatively less common within the general population, such as ‘goiter’ (*freq*=0.029), the composition of symptoms becomes increasingly similar to the representation of Graves’ disease, and the similarity with the other three autoimmune conditions begins to diverge. These patterns mirror how individual symptoms may not provide sufficient information to assess the probability of a specific condition, but as a patient accumulates more symptoms over time, the aggregation of these symptoms can stratify patients by disease risk.

### Clinical knowledge embeddings stratify patients by disease risk and severity

We next investigate how *ClinVec* can be utilized in the context of patient outcome prediction, including disease progression and disease risk. Using only information derived from *ClinVec*, we compute a disease-specific risk score for various conditions and the relative disease risk for each patient. Analogous to polygenic risk score frameworks^99^, we compute a phenotype risk score^100^ by scanning a patient’s EHR and aggregating the embeddings across conditions with the highest similarity to the disease of interest (Methods). We validated our approach using retrospective EHR data from 4.57 million individuals^101^ at CHS and specifically focused on patients listed in the CHS chronic registry, a registry maintained by CHS that monitors individuals with specific chronic conditions^102^. We examine three clinical conditions with the largest sample sizes within the chronic registry: chronic kidney disease (CKD), chronic obstructive pulmonary disease (COPD), and heart failure.

For each condition, we constructed a matched case-control cohort (Supplementary Table S6, Supplementary Figure S5) and used 5 years of clinical history to compute a relative risk score for each patient. For a target disease (here represented by an ICD10CM embedding), we identify embeddings (features) most relevant to the disease by selecting all codes within a given radius in the embedding space (see Methods). Each patient’s score is computed as a weighted sum of these features, where the weight is calculated as the cosine similarity between the feature embedding and target disease embedding. In practice, the size of the radius (k-neighbors) can be selected by comparing a validation patient dataset, similar to choosing the optimal performing p-value threshold in PRS^103^. This approach eliminates the need to pre-specify a list of features useful for diseases with heterogeneous presentations and highly variable symptoms, such as many chronic conditions.

Quantifying patients’ disease risk according to their risk score percentile of the total score distribution, we observe the separation of patients according to disease status across all three condi tions, particularly at the extreme end of the risk distribution (Figure 5). Within the group of individuals in the 10^th^ percentile of the risk score distribution for CKD, there is a disease prevalence of 6.2%. For patients above the 99^th^ percentile, the disease prevalence increases to 23.4%. Top identified clinical code embeddings are consistent with each disease’s known comorbidities, lab test results, and medication usage. For example, among the top features of chronic kidney disease is *Diabetes mellitus*, which is a well-documented risk factor for the disease, and *Proteinuria*, which can be an early disease indicator. Although we assess the population-level correlation between disease prevalence and risk score percentile, these results suggest opportunities to leverage clinical knowledge embeddings for patient monitoring in a way that is independent of any training dataset.

**Figure 5:**
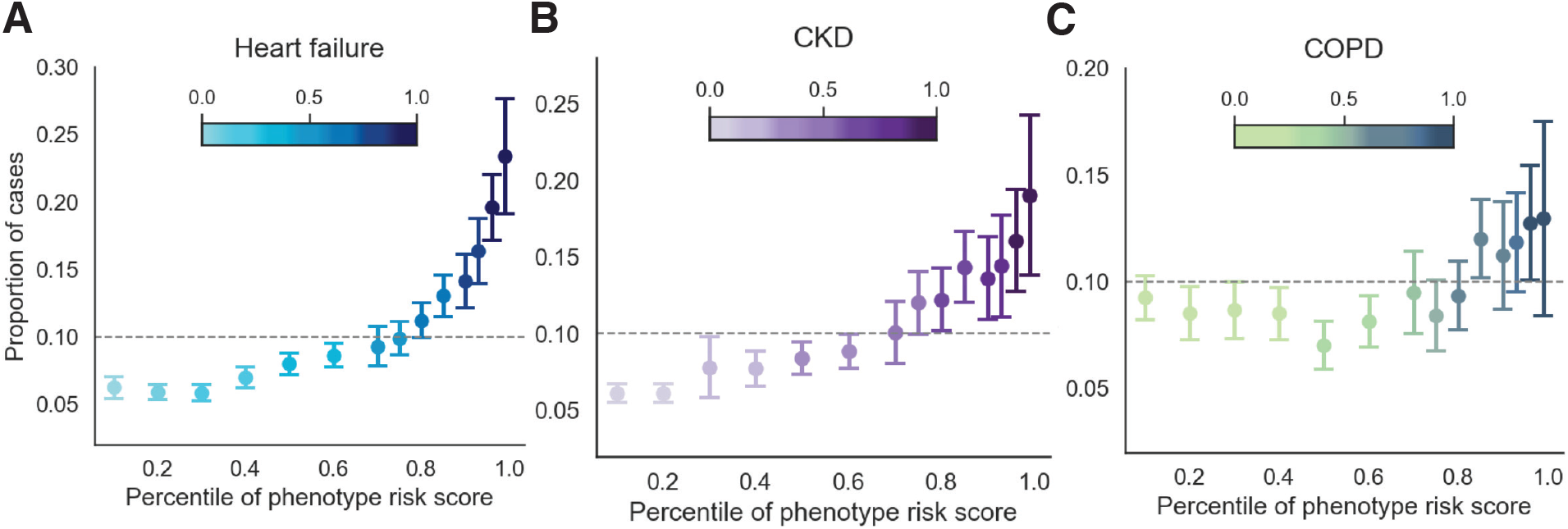
Clinical code embedding risk scores correlate with disease prevalence and severity. **(A-C)** We provide scatter plots showing the risk score percentiles of each disease versus the disease prevalence within each bin. Dots are shaded according to disease prevalence. Columns are divided by diseases: heart failure, CKD, and COPD.

Given the marked stratification by disease status, we evaluate whether the computed risk scores could provide disease subtyping information. Intuitively, suppose individuals acquire a greater number of highly relevant clinical conditions. In that case, this trend may indicate a later disease stage and increased mortality rates. To prevent confounding from disease-specific effects with age and sex, we restrict the assessment to individuals of the same sex and diagnosed at a similar age (Methods). For each of the three conditions, we compute the mode age and sex and use this as inclusion criteria for the survival analysis. Re-calibrating score percentiles based on the final set of patients, we observe a striking stratification by 6-year survival rate (Supplementary Figure S6). For CKD (age=72 sex=Male), individuals within the top quintile of this score distribution for CKD had a relative 42.6% lower 6-year survival compared to those in the bottom quartile (top-quartile: 56.8.3%, *bottom-quartile*: 76.5%). This trend is particularly remarkable considering the risk scores were inferred without any institution-specific, disease-specific, or patient level information during training.

### *ClinVec* provides structured clinical knowledge to improve LLM performance

Clinical embeddings provide a structured approach to incorporate domain-specific knowledge into foundation models such as LLMs. One common strategy is knowledge injection, where embeddings from external knowledge sources are appended to the LLM input. Recent studies show that this approach improves performance on tasks such as medical question answering by grounding LLM outputs in a clinically relevant context^104–107^. We adapt MMed-Llama^106^, an open-source LLM composed of 8 billion parameters. To test whether *ClinVec* embeddings improve LLM performance beyond what can be achieved with domain-specific training data alone, we evaluated our knowledge token injection approach using an LLM already trained on medical content. We used parameter-efficient fine-tuning to integrate 42,883 clinical knowledge tokens into the LLM, where each token corresponded to a *ClinVec* embedding of a UMLS concept (Supplementary Figure S7). This approach allows domain adaptation without retraining the entire model or requiring a labeled clinical dataset. It provides a resource-efficient strategy for incorporating structured clinical knowledge into LLMs.

To assess the impact of clinical knowledge embeddings on LLM performance, we used the MedQA^1–4^ dataset, which contains questions from the US Medical Licensing Exams (USMLE).

We applied the UMLS MetaMap API to map question text to UMLS concepts, then retrieved the corresponding *ClinVec* embeddings and appended them to the tokenized prompt (see Methods). This approach incorporates structured, clinically grounded knowledge without retraining the underlying LLM. Instead, *ClinVec* supports retrieval-augmented generation, where embeddings relevant to the input are used to guide the LLM response.

Assessing performance with a held-out test set of MedQA (N=1,247 questions), we find a 10.3% increase in question-answering accuracy when incorporating the clinical knowledge embeddings (acc=0.64) compared to question answering without it (acc=0.58). We observe the largest performance gains when questions are short, likely because the language model has limited contextual information to generate accurate answers. Adding just one or two clinical knowledge embeddings leads to an average improvement of 11.7%, suggesting that the relationship information encoded in these embeddings helps compensate for missing context. Performance continues to improve as more tokens are added, reaching a peak at around 9 to 10 tokens. Beyond this point, additional embeddings introduce less relevant or less specific information, which reduces overall accuracy.

**Figure 5:**
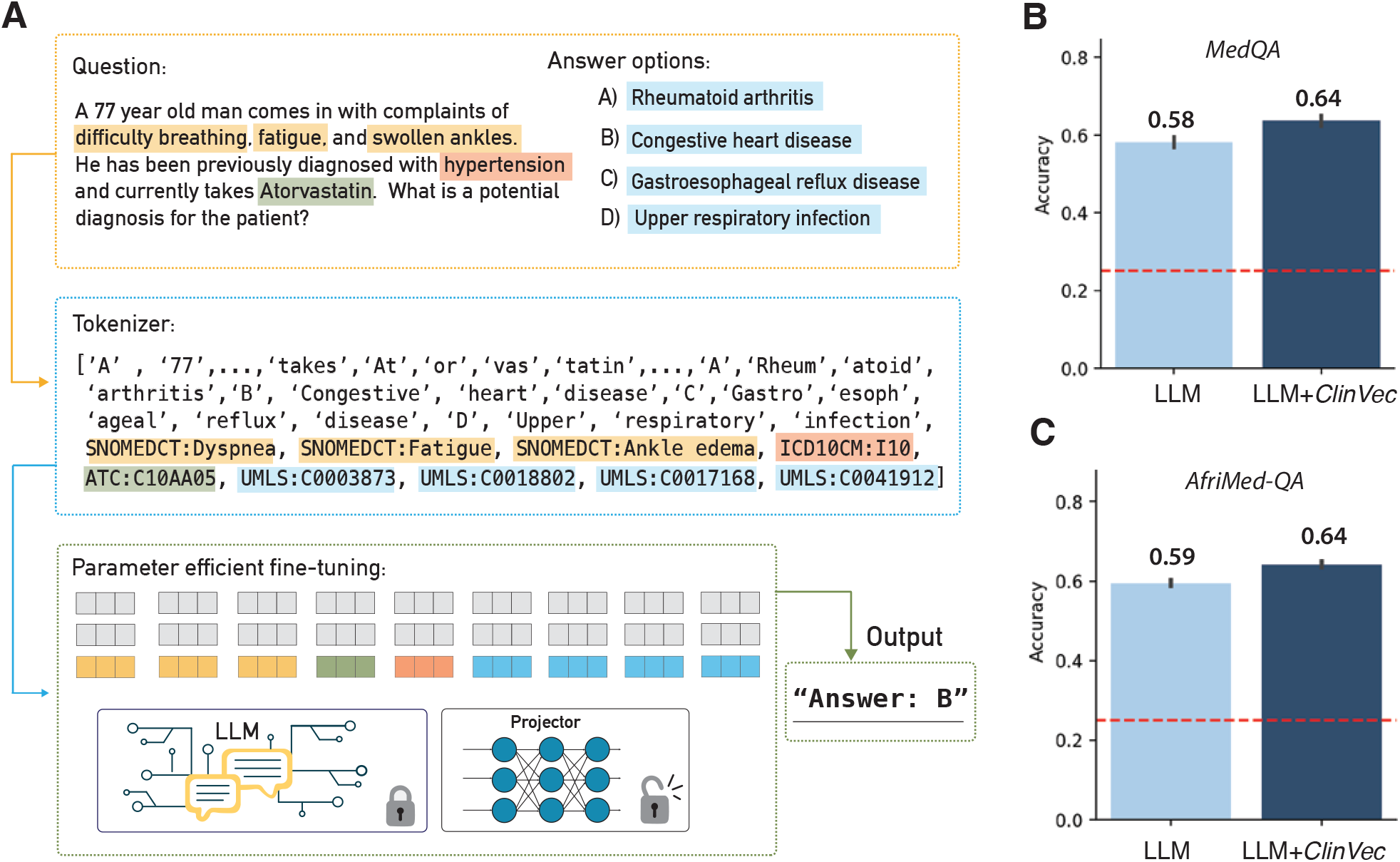
Integrating *ClinVec* with large language models improves medical question-answering. **(A)** Overview diagram of question-answering with knowledge injection using *ClinVec*. **(B)** We report the accuracy of N=1,247 medical questions in the MedQA dataset. The red line denotes random performance with four candidate answer options. **(C)** Accuracy of N=3,724 multiple-choice questions from AfriMed-QA dataset. The red line de notes baseline performance with five answer options.

To assess generalizability across different clinical scenarios, we also examined the recently released AfriMed-QA dataset^109^, which consists of 3,724 multiple-choice questions collected from healthcare settings across 16 African countries. Since the dataset was released in early 2025, there is no data leakage between the training and fine-tuning datasets. We observe a similar magnitude of performance improvement where we observe accuracy of 0.64 with the addition of clinical knowledge embeddings compared to 0.59 without—a 8.5% increase. Breaking down performance by clinical specialty (a label provided in the original dataset), we identify the specialties with the greatest performance gains, including hematology (+37.0%) and ophthalmology (+20.3%). Given the specificity of these fields, the addition of clinical knowledge yields the most substantial improvements in question-answering performance. Hematology shows the largest gains, likely due to the distinct disease presentations encountered in African settings that are underrepresented in the training data of most large language models. For instance, one correctly answered question concerns antiretroviral treatment regimens for HIV-associated anemia, a clinical scenario more common in regions with high HIV prevalence. This example illustrates how anchoring model responses in structured, domain-specific knowledge from *ClinVec* helps address regional variation and improves accuracy in specialized medical contexts. We observe consistent performance improvements across both U.S.- and Africa-based clinical questions, suggesting that *ClinVec* supports generalizable medical reasoning.

## Discussion

We introduce *ClinGraph*, a clinical knowledge graph, and *ClinVec*, a corresponding set of 153,166 clinical vocabulary embeddings, that together translate structured clinical information into a machine-readable format for use in AI models. These embeddings capture patterns among clinical codes that reflect human anatomy, disease presentation, and healthcare organization. They encode semantic relationships that support the combination and comparison of clinical concepts. We demonstrate that the embeddings enable patient-level prediction across diverse condi tions without using patient data during training. Evaluations by inter-institutional panels of clinicians from the United States and Israel confirm that the embeddings align with current clinical knowledge and practice. This resource offers a generalizable framework for sharing clinical knowledge and developing transferable, knowledge-grounded AI models.

Clinical code embeddings offer a way to represent structured clinical knowledge for use in AI models. By encoding relationships among diagnoses, procedures, medications, and laboratory tests, the embeddings create a knowledge-grounded representation of clinical data that can be used in downstream machine learning applications. These representations are well-suited to serve as inputs to foundation models, large-scale models pre-trained on unlabeled data and adapted to specific tasks through fine-tuning with labeled datasets^110^. The foundation model approach has shown success in natural language processing and computer vision, supported in part by the availability of publicly shared models such as DeepSeek^111^ and LLaMA^112^. Our resource can be used in retrieval-augmented generation^113^ systems where embeddings serve as a structured knowledge base. In this setting, relevant embeddings can be retrieved based on a user prompt to provide clinical context and support more accurate and informed model outputs^41^.

While this resource offers a foundation for advancing precision medicine, it has several limitations. First, the current knowledge representations do not model uncertainty. Clinical knowledge is derived from heterogeneous sources, and the frequency or relevance of specific observations can vary across populations. For example, while disease guidelines may list several symptoms, their prevalence and clinical importance often differ between individuals. Second, the quality of the embeddings depends on the accuracy of the underlying KG. Errors such as incorrect or in complete edges can introduce noise and distort the learned representations. As clinical knowledge evolves, the graph must be regularly updated to reflect current clinical standards and practice. Third, the embeddings do not incorporate contextual information, such as age or sex, which can influence the presentation and interpretation of clinical concepts. For example, the clinical presentation of a heart attack can vary significantly between males and females^114^, but this delineation is not considered in the construction of the embeddings.

*ClinVec* embeddings cover common and chronic diseases, with limited representation of genetic disorders and Mendelian conditions. Future work will incorporate codes from phecodeX^115^, an updated vocabulary that includes chromosomal anomalies, as well as resources focused on Mendelian diseases, such as the Online Mendelian Inheritance in Man (OMIM)^116^ and Human Pheno type Ontology (HPO)^74^. We also plan to integrate databases that capture biological mechanisms, including those involving genes and proteins, to improve the representation of disease with complex or poorly defined clinical features.

*ClinGraph* consolidates fragmented coding systems into a unified representation of clinical knowledge, addressing interoperability challenges while protecting patient privacy. The embeddings are designed to capture clinical knowledge that generalizes across care settings and aligns with established medical practices. Unlike datasets that only reflect recorded health states, *Clin Vec* embeddings incorporate information about clinical guidelines, mechanistic associations, and disease-treatment pathways. Because the embeddings are derived from structured vocabularies and not patient-level data, they can be shared across institutions without risk of exposing identifiable information. Unlike models pre-trained on patient records, which may inadvertently retain or memorize sensitive information^117,118^, *ClinVec* embeddings can be shared across healthcare systems without risking patient privacy. This approach also reduces dependence on specific patient populations and supports diverse clinical applications. By embedding structured clinical knowledge into AI models, this resource helps ensure that predictions are grounded in established medical understanding^119^. Resources such as unified clinical vocabulary embeddings can strengthen the reliability and generalizability of AI models.

## Methods

### Standardized medical vocabularies

We use standardized medical vocabularies to construct all clinical concepts in *ClinGraph* and *ClinVec*, supporting broad applicability across clinical use cases: Unified Medical Language System (UMLS), International Classification of Diseases (ICD)^120^, Anatomical Therapeutic Chemical (ATC) Classification^121^, RxNorm^122^, Systemized Nomenclature of Medicine – Clinical Terms (SNOMED CT)^123^, Current Procedural Terminology (CPT)^124^, Logical Observation Iden tifiers Names and Codes (LOINC)^125^, and phecodes^126^. Here, we define a clinical code as an en try within a medical vocabulary with a unique identifier.

We begin by integrating the clinical codes and relationships defined in the UMLS vocabulary. We incorporate all UMLS CUIs (concept unique identifier) with a source vocabulary in ICD10CM, SNOMED CT, LOINC, RxNorm, ATC, or CPT (N=42,883 nodes). Next, we inte grate all ICD10CM codes up to 2 decimal places (N=34,842) and all ICD9CM codes. We also add information from the phecode coding system which represents clinical phenotypes as phe codes, formed by aggregating groups of diagnostic codes (i.e., ICD codes) into clinical sub groupings (N=1,817) ^127–129^. Phecodes were developed to facilitate the secondary use of EHRs for biomedical research rather than for patient monitoring or hospital administration. Numbers after the decimal point reflect a hierarchical structure similar to the ICD code hierarchy. Under each leaf phecode is the set of corresponding ICD-9/10 codes where each phecode has between 1 – 20+ associated ICD-9/10 codes. Next, we add nodes representing LOINC Part codes, which are standardized attribute values used to construct the LOINC codes that specify a given laboratory test. We restrict our inclusion to only LOINC Part component codes which refer to the substance being tested (N=14,399). We also incorporate all CPT codes (N=4,377), ATC codes (levels 3-5: N=5,179), and RxNorm drug groups (N=10,130).

Next, we integrate a subset of SNOMED CT codes as nodes in *ClinGraph*. Because the SNOMED CT vocabulary includes many codes (>350,000), we wanted to restrict the inclusion to the most clinically relevant codes. We begin by selecting all SNOMED CT codes from the CORE (Clinical Observations Recording and Encoding) Subset, a set of most frequently used codes provided by UMLS constructed to maximize data interoperability across institutions. Next, we include SNOMED CT codes from the Convergent Medical Terminology subset diagnosis list, a set of core terms originally curated by Kaiser Permanente and now used across numerous healthcare systems.

### Relationships between clinical concepts

To construct edges between pairs of clinical concepts (nodes), we first incorporate relationships defined by the PheMap database, an existing knowledge base of biomedical knowledge that defines a set of relationships between the codes and various clinical vocabularies^83^. This database offers relationship pairs linking phecodes and clinical codes from other medical vocabularies, including RxNorm, CPT, LOINC, and SNOMED CT. Relationships defined in PheMap were determined through text mining and natural language processing of open-source biomedical literature. These relationships can be interpreted as a phenotype being “associated with” a given clini cal concept. To construct relationships between different medication codes, we use ATCPROD, a product-level mapping of RxNorm to ATC-4 classes. Edges between ICD9CM and ICD10CM codes are derived from the General equivalence mappings (GEM), the official crosswalk mapping diagnoses codes to and from ICD9CM and ICD10CM.

### Constructing ClinGraph

To efficiently represent clinical knowledge in a format optimized for machine learning, we formally organize the clinical codes and relationships, as described above, into a heterogenous knowledge graph^130^ (KG) called *ClinGraph*. We draw an undirected edge between two clinical codes if they have a pairwise relationship as described above in the clinical vocabulary aggregation process. Multiple edges are drawn if two nodes have a relationship defined multiple times (e.g., a relationship between a phecode and ICD-9CM code derived from the phecode hierarchy and the PheMap knowledge base). The final graph comprises 153,166 nodes across seven node types and 2,873,025 edges, and the distribution of nodes and edges can be found in Supplementary Table S1, S2.

We model 4 distinct node types: diagnosis codes (ICD9CM, ICD10CM, phecodes), medications (ATC, RxNorm), laboratory tests/procedures (LNC, CPT), and clinical concepts/vocabularies (SNOMED CT, UMLS). By grouping similar vocabularies (e.g., RxNorm and ATC), we allow for information sharing across similar nodes. However, modeling distinct edge types would require the harmonization of relationship types across databases. For databases without defined edge type semantics, such as PheMap, this harmonization process would likely require additional resources and clinical expertise. For this reason, we only model distinct node types and leave the implementation of edge-type specific embeddings as future work.

### Heterogenous graph transformer

#### Graph notation

A heterogeneous graph, 𝒢 = (*V*, ℰ), is defined by a set of nodes *V* and edges ℰ. We define a mapping function *f*_*v*_:*V* → 𝒯^*v*^ where each node *v* ∈ *V* has an assigned node type *f*_*v*_(*V*) ∈ 𝒯^*v*^. Here, the node types represent different clinical vocabularies. For a given graph, 𝒢, we denote the graph neural network, ℋ_𝒢_, as a function that maps a vector of features for a node μ, written as *x*_*u*_ ∈ ℝ^*d*^, to a vector of real numbers representing the latent space:

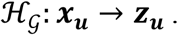

Here, *d* is the dimension of the input vector. This resulting mapped vector (also called an *embed ding*) is denoted by ***z*** ∈ ℝ^*d*^. For convenience of notation, we assume that the input and output dimensions are equal. We provide a table with relevant notation and indicate whether they are an inferred parameter within the model.

**Table 1:**
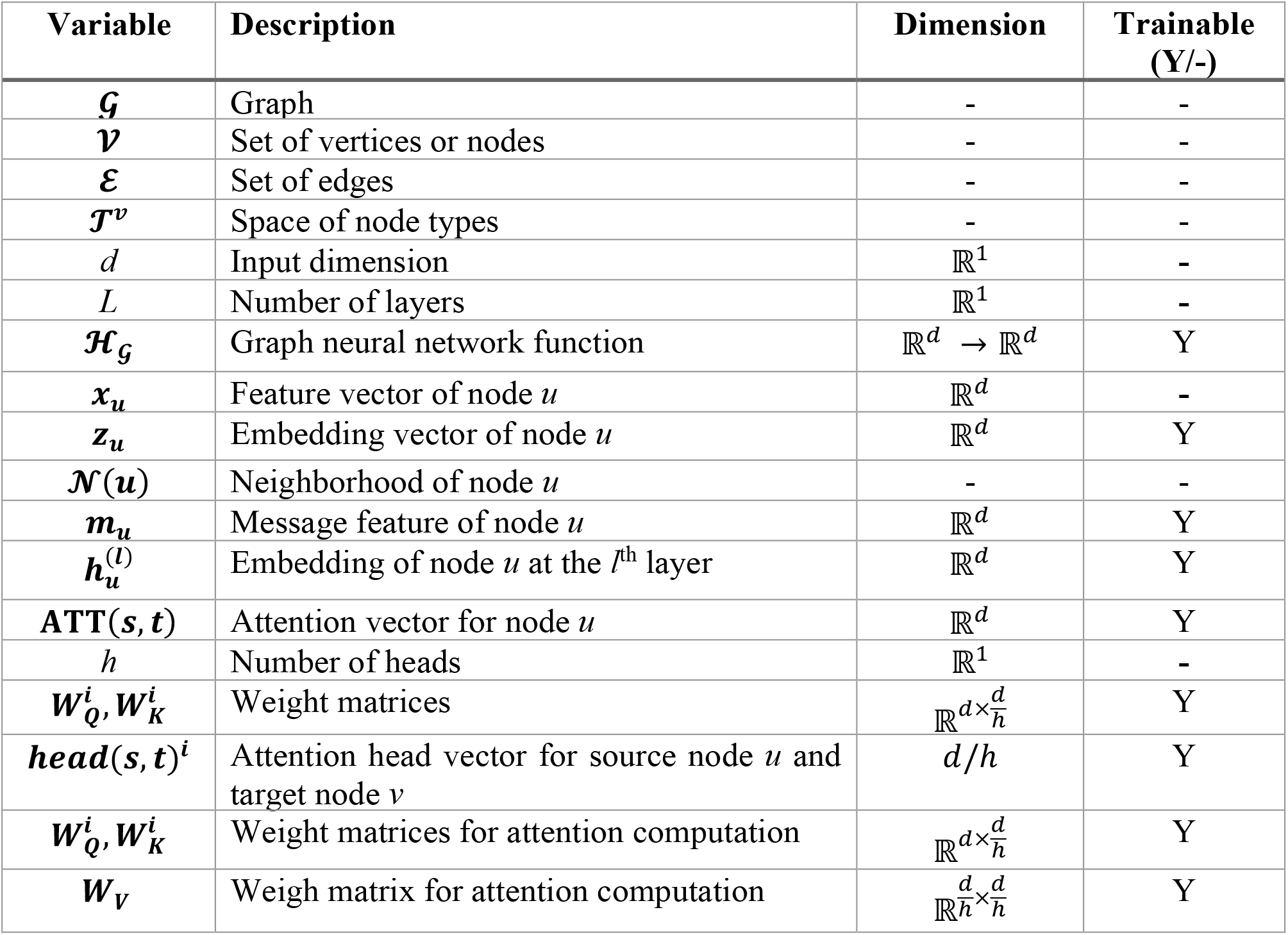
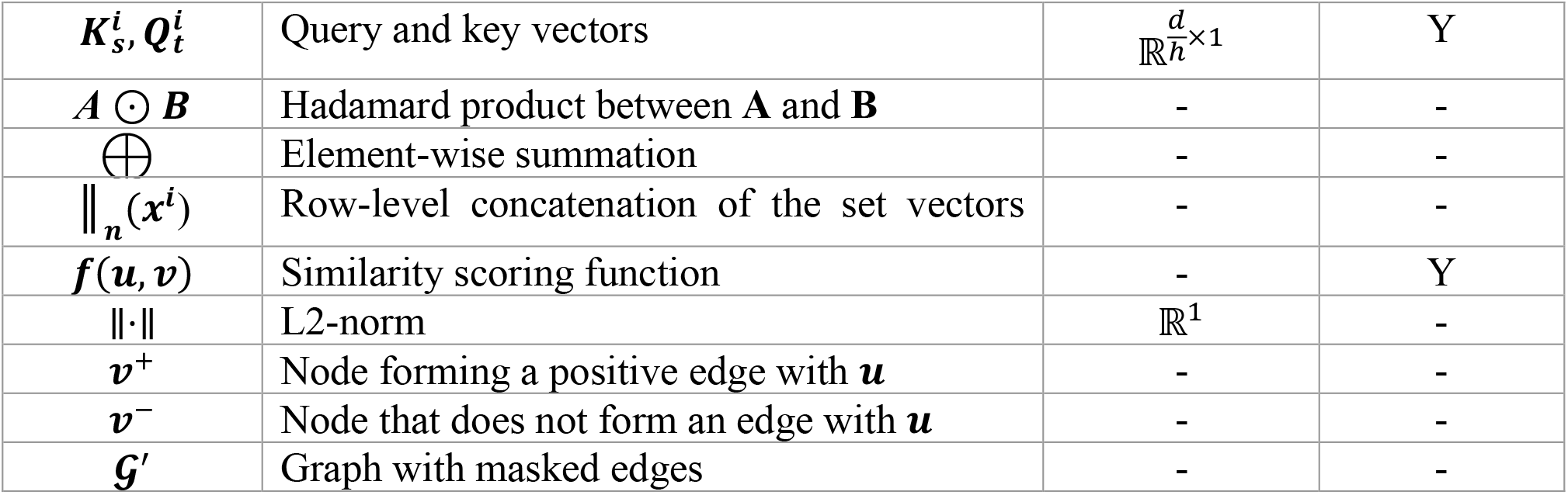
Summary of notation. We provide a summary of the variables used in deriving the graph transformer network model, the dimension of each variable, and whether it is a learned or fixed parameter. Rows are sorted according to the order in which each variable is introduced.

#### Overview

We model the architecture of our graph neural network (GNN) after the heterogeneous graph transformer (HGT)^86^. In this work, we briefly describe the key model steps. The complete deri vation of HGT is detailed in previous work^86^.

Learning ℋ_𝒢_ involves aggregating information across the graph structure defined by 𝒢. For a target node *t*, we define its neighborhood, 𝒩 (*t*), as the set of nodes with a direct connection or edge with *t*. Information is propagated through the graph by gathering information from the neighborhood of *t*, which is aggregated into a message feature ***m***_*t*_. The embedding of *t* can be updated with the resulting message feature, 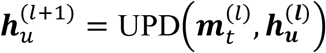 where 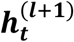 is the embed ding of node *t* at the *l+1* layer and UPD(·) refers to an update function that integrates messages with the current node embedding. Note that the input of the *l+1* layer is the output of the previ-ous layer, and by stacking L layers 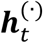, we represent the final HGT as a composition of these functions across all layers:

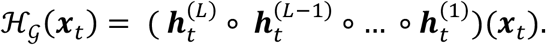

The input of the model is the set of node features, ***x***_*t*_ ∈ ℝ^*d*^ for all nodes in 𝒢 which is then used to initialize the 0^*th*^ layer embedding of each node 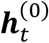. In practice, each of the node features is initialized using Xavier noise. The embedding represents the relational information learned from the knowledge graph without additional information from the node features.

### Multi-head attention mechanism

The HGT uses multi-head attention, which is a key feature of transformer architecture^87^. Multi head attention utilizes multiple attention mechanisms, or heads, in parallel to tend to different parts of the input, where each head has its own set of learnable weights (parameters). Updating a node’s embedding involves aggregating messages across all *h* heads and weighting each mes sage by its attention score. This mechanism essentially up- and down-weights messages based on their importance so that only necessary information is propagated to the target node. Although the original HGT architecture allows the modeling of different relation types within heterogene ous graphs, we treat all nodes and edges as a single type during inference to maximize infor mation sharing across the knowledge graph.

For a target node *t*, a single attention head is a series of linear projections of the embedding from the previous layer, normalized by the embedding dimension. Here, a linear projection for an in-put ***x*** ∈ ℝ^*d*^, is defined as an affine linear transformation, ***y*** = ***W***^***t***^ ***x*** + ***b*** where ***b*** is a bias term and ***W*** is a learnable weight matrix. For convenience, we drop the bias term in the following derivations, but in practice, the parameter corresponding to the bias term is absorbed into the estimation of ***W***.

The series of linear projections mirror the standard query, key, and value vectors used in the standard transformer architecture. The attention score vector corresponding to the edge between source node *s* and target node *t*, represented as **ATT**(*s*,*t*), is computed as the concatenation of each head’s output:

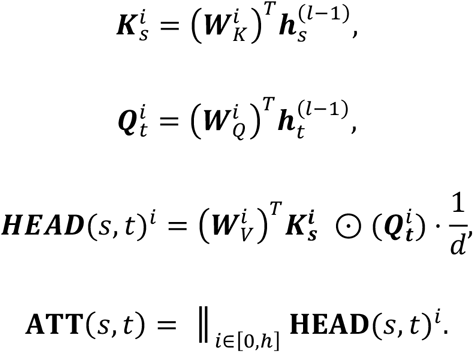

Here, the operator “***A*** ⊙ ***B****”* represents the elementwise or Hadamard product between ***A*** and ***B***, and the operator “║_*n*_(***x***^*i*^)” represents the row-level concatenation of the given set of vectors {***x***^1^, …, ***x***^*n*^}.

The input vector is 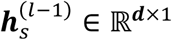, and the learnable weight matrices are 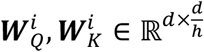, and 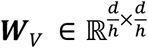 where 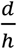 is the input dimension divided by the number of heads. For these weight matrices, the subscripts refer to the *query, key*, and *value* elements in the transformer. The result ing query and key vectors follow as 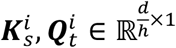. The resulting attention vectors from each head and final multi-head attention vector are 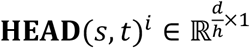 and **ATT**(*s*,*t*) ∈ ℝ^*d*×1^. And to ensure the attention scores for the target node *t* sum to 1, a *Softmax*(·) is applied across all neighbors of *t*.

#### Message passing

For target node *t*, we first compute the message vectors across all heads for each neighbor of *t*. This message is calculated as a linear projection of the embedding for source node *s* from the prior layer multiplied by a weight matrix (***W***_***T***_). Each head maintains its linear projection matrix but ***W***_***T***_ is shared across all attention heads.

The final message passed from *s* → *t* is a concatenation of messages produced from each of the *h* heads. This resulting message vector is then multiplied by the multi-head attention vector. The final updated embedding 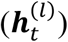 is computed by aggregating across all neighbors of the node *t* through an elementwise sum. This is followed by a linear projection (***W***_*N*_), which creates separate projection for each node type. We need a single matrix because we only model a single node type. This is then followed by a residual connection and passed through an activation function:

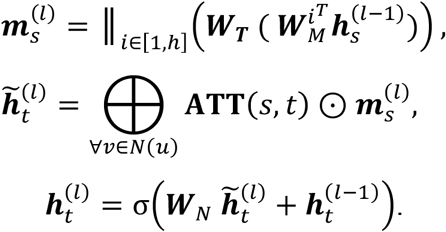

The learnable weight matrices are 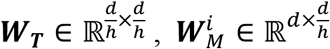, and ***W***_*N*_ ∈ ℝ^*d*×*d*^. The message vector is 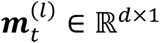. The operator ⨁ represents the elementwise summation. Training the final model involves optimizing the set of learnable weight matrices, 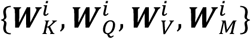 across *h* heads and finding {***W***_*T*_, ***W***_*N*_} that minimizes the loss function ℒ.

### Self-supervised learning

For a given node *u*, our goal is to generate an embedding ***z***_*u*_ that quantifies the topological infor mation of the graph w.r.t *u*. The learning objective can be framed as a link prediction problem, where given two nodes *u* and *v*, we want to train a function to predict whether an edge exists between these two nodes. This function can be described by,

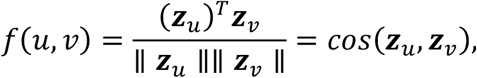

where ***z***_*u*_ = ℋ (*u*), and ℋ (·) represents the trained GNN function that provides the node embeddings. Note that because edges are undirected and the scoring function is symmetric, *f*(*u, v*) = *f* (*v, u*).

Optimizing this above scoring functions means solving for the parameters of ℋ(·). This can be performed using contrastive learning. We mask a random subset of edges to create a modified version of the graph, 𝒢′. Then, given a source node *u*, we define a node where *v*^+^ ∈ 𝒩 (*u*) represents a target node that forms an edge with *u* in the original graph 𝒢. We denote *v*^−^ ∉ 𝒩 (*u*) as a node that does not form an edge with *u* in 𝒢 and denote the set of these negative examples as 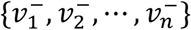.

The function *f*(*u, v*) is trained on 𝒢^′^ to estimate whether a source node *u* forms an edge with target node *v* in the original graph 𝒢. We can quantify the probability of an edge existing between *u* and *v*^+^ with negative edges 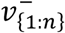 as the probability,

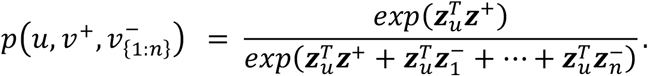

Given a dataset of *N* source nodes, each with a corresponding set of candidate nodes as described above, the negative log-likelihood over the dataset is equivalent to the cross-entropy loss where the true “class” always corresponds to the positive node *v*^+^. This type of contrastive loss function is well-established^88,131–133^ and is formulated as follows:

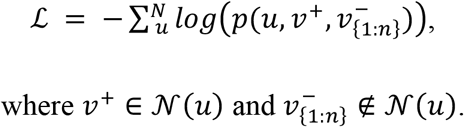

### Node type-aware sampling of positive and negative edges

The contrastive learning requires specifying positive and negative edges during training, where a positive edge is defined as a pair of nodes with an existing edge in *ClinGraph*, and a negative edge is a pair of nodes that are not connected by an edge. To generate a training tuple comprised of a single positive edge and a set of corresponding negative edges, we randomly sample an existing edge within *ClinGraph* denoted as (*u, v*) and 𝒯^*v*^ (*v*) is the node type of target node *v*. To create a negative sample, we select a node *r* such that 𝒯^*v*^ (*r*) = 𝒯^*v*^ (*v*) and *r* ∉ 𝒩 (*u*).

### Stochastic mini-batch training with neighbor sampling

Because the topology and size of the GNN are determined by the KG, storing the entire network and its hidden states onto a single GPU is impossible. Although training with mini-batches alleviates some of this issue, the computation graph (the set of nodes involved in message passing in each iteration) for a GNN with *L* layers often still includes millions of parameters. To circumvent this, we use node neighbor sampling to select only a subset of nodes to include at each layer when computing a node’s message vector. Specifically, we take a sample n of neighbors at each k-hop neighborhood at each gradient descent step when computing messages during optimization. Node types with a smaller set of nodes *ClinGraph* are upweighted to ensure they are represented in the sampling. The number of samples at each layer is a hyperparameter selected through hyperparameter tuning. The final training was performed on an H100 GPU for ten epochs, during which we trained on all 2,873,025 edges of the KG.

### Hyperparameter tuning

We perform hyperparameter tuning in 2 steps: first for the model architecture and then for the parameters involved in the contrastive learning objective. We select a random subset of 500,000 edges for training and 100,000 for validation from *ClinGraph*. We perform hyperparameter tuning using Raytune where we fix the learning rate (1e-4), number of negative samples (10), batch size (100), and sampling neighborhood (10) and vary the following parameters: number of layers (2, 3, 4), input feature dimension (128, 256, 512), output embedding dimension (128, 256, 512), head dimension (128, 256, 512), number of heads (2, 3, 4), and dropout (0, 0.1, 0.2). Performance is assessed by measuring the prediction accuracy on the set of 100,000 validation edges and 100,000 node pairs that are not linked in the KG (negative edges). The optimal set of GNN parameters is 4 layers, 128 input dimensions, 128 output embedding dimensions, 512-dimension head size, three heads, and a dropout of 0.0.

We repeat the hyperparameter selection process by fixing the parameters selected above and vary the following: learning rate (1e-3, 1e-4, 1e-5), neighborhood sampling (5, 10, 20), and number of negative samples (3, 5, 10). The final hyperparameter set selected is a learning rate of 1e-5, neighborhood sampling of 20 nodes, and three negative samples. All experiments were performed using the Ray Tune software (https://docs.ray.io/en/latest/tune/index.html).

### Clinical evaluation study

A key transferability aspect is demonstrating that the inferred clinical knowledge embeddings capture medical insights that align with established clinical consensus. Although the knowledge graph construction was based on mined biomedical literature, the inferred embeddings and latent space offer insight into relationships between codes beyond the direct edges in the KG. To validate this, we perform a human pilot evaluation with groups of clinical experts from Brigham Women’s (N=1), Mass General (N=1), Boston Children’s Hospital (N=1), and CHS (N=3).

To systematically assess this, we evaluate the association between different diseases and SNOMEDCT US codes. We restrict the set of SNOMED codes to the Clinical Observations Recordings and Encoding (CORE) Problem List Subset of SNOMED CT^48^. This subset of codes has been identified as most informative in documenting and encoding clinical information. We survey clinicians to select 90 disease phenotypes (phecodes) that broadly capture major disease areas and are clinically relevant for potential clinical AI applications. For each disease, we compute the cosine similarity between each SNOMED CT code that does not already form an edge with the target phecode. We select the top 20 SNOMED CT codes for evaluation. A control set is constructed by randomly selecting 20 SNOMED CT codes that are not in the top 20 nor already form an edge in the KG. To prevent biases due to differences in the frequency of symptoms or high degree and low degree within *ClinGraph*, we match control SNOMED CT codes based on the case code’s node degree.

Clinicians were provided with shuffled disease lists of case and control codes. For a given disease, a single clinician evaluated the full list of codes. They were provided a brief background of SNOMED CT codes and a scoring rubric where they were asked to evaluate each SNOMED CT code according to how related each code is to the given disease phenotype. We utilize a Likert scale^134^ based on scores 1 – 5: ‘unrelated’ (0) and ‘ highly related’ (5). If a code is too broad or non-specific (e.g. ‘Clinical evaluation’), these are assigned a 0.

### Visualizing the latent space

To visualize the *ClinVec* embedding space, we perform UMAP^135^ directly on the embedding vectors and plot the first two dimensions. When performing dimensionality reduction, we use phe codes, ICD10CM, ATC, LOINC, CPT, and SNOMED CT embeddings. Phecode categorizations are pre-defined from the phecode v1.2 mapping^136^. To visualize the latent space of phecodes within the digestive system, we performed UMAP only on the subset of ICD10CM embeddings within the ‘Digestive’ category. We highlight ICD10CM codes broadly corresponding to mouth/teeth conditions (phecodes 520-530), functional digestive disorders (560-570), and biliary tract disorders (570-580). To highlight embeddings related to specific diseases, we compute the cosine similarity between an ICD10CM embedding (representing a disease) and each embedding within the latent space. We visualize this relationship by shading each embedding within the UMAP space according to the cosine similarity. We highlight relevant codes for lupus and type 1 diabetes to provide additional interpretation of the latent space map. This information was derived from the guidelines provided by the National Resource Center on Lupus^137^ and the Centers for Disease Control and Prevention^138^.

#### Comparing embeddings from PrimeKG and SNOMED-CT KG

We generate embeddings for PrimeKG and SNOMED-CT KG using the same graph transformer architecture and hyperparameters as *ClinGraph*. For PrimeKG, we modeled 10 node types as defined by the KG. For SNOMED-CT KG, we only used a single node type (i.e. homogenous graph) since all nodes represent SNOMED CT codes. We trained both models for 5 epochs and generated embeddings of size 128 for all SNOMED CT codes in SNOMED-CT KG and all diseases (MONDO, HPO nodes) and drugs (DrugBank nodes) for PrimeKG. Clustering metrics, including silhouette score and adjusted mutual information, were computed according to ATC level 1 and phecode category.

Because neither KG have ICD or ATC codes, we mapped the nodes as follows. For SNOMED CT KG, we mapped each SNOMED CT code to its UMLS CUI and then to its corresponding ICD10CM code if present. If a UMLS CUI does not have a corresponding source ICD10CM code, we removed the node embedding from the cluster calculation. To map SNOMED CT to ATC, we first mapped SNOMED CT codes to its UMLS CUI. We then used UMLS to identify UMLS CUI codes with ‘ingredient-of’ relationships that describe the active ingredient (represented as ATC) present in a given UMLS CUI drug. SNOMED CT codes that did not have a mapped ATC ingredient were removed from the clustering calculation. Clustering metrics were computed using all 128 dimensions from each set of embeddings, and cluster visualizations were produced using UMAP.

#### Zero-shot embedding retrieval tasks

We downloaded embeddings from snomed2vec (https://github.com/KhushbuAgarwal/Snomed2Vec) and cui2vec (https://github.com/beaman-drew/cui2vec) directly from the original publications. We formulated a set of 12 retrieval tasks using well-established disease-symptom and disease-indication lists (Supplementary Table S8). Top ranked neighbors are determined using cosine similarity between embeddings. We report each model’s performance using ‘hits @ k’ which measures the proportion of diseases where a relevant symptom/drug is ranked among the top k items within the embedding space.

### Disease embedding arithmetic

#### Constructing disease symptom embeddings

To demonstrate the syntactic and semantic properties of *ClinVec*, we use vector-level arithmetic to aggregate together disease symptoms to form a ‘disease symptom embedding’. Aggregation is performed using sum pooling, but any vector-level aggregation method could be used in practice. For a given disease, we retrieve the list of symptoms from the Mayo Clinic Disease Description Guidelines^139^. A clinician manually translates each symptom into its corresponding ICD10CM code. We provide these translations for all nine diseases discussed within this analysis (Supplementary Table S5). To demonstrate the trend of increased similarity between a disease embed ding and the disease symptom embedding, we order symptoms from highest to lowest prevalence. We approximate population prevalence using a random sample of 200,000 patients from the CHS database.

Disease symptom embeddings are first initialized with a vector of zeros. We then add the embeddings from all symptoms (represented by an ICD10CM embedding). For visualization, we compute the cosine similarity between the disease symptom embedding at each step and the target disease embedding, represented using an ICD10CM embedding. Across diseases, we see a broad trend of increased similarity with the disease embedding as more symptoms are aggregated.

#### Robustness analysis

To assess the robustness and generalizability of these trends, we perform the same analysis under various conditions while perturbing different variables in the framework. This includes analyses assessing the uncertainty in selecting disease symptoms, incorrect disease specification, and the impact of random symptoms. First, due to the noise and uncertainty associated with using diagnosis codes as proxies for disease symptoms^13^, we tried to model real-world circumstances by varying the ICD10CM code used to represent each symptom. We selected a set of candidate ICD10CM codes for each symptom by taking the original code from the clinician review and selecting all ICD10CM codes with the same parent code by removing one decimal place. For example, for code ‘786.2’ (Cough), the candidate set would include ‘786.1’ (Stridor), ‘786.0’ (Dyspnea and respiratory abnormalities), etc. Then, when selecting each symptom for aggregation, a random code from the candidate set is selected. This process is repeated 10 times, yielding similarity curves constructed from 10 different disease symptoms.

Next, we wanted to simulate when the disease of interest is mis-specified compared to the symptom list. This reflects situations where the symptoms do not describe the target disease of interest but, instead, a closely related condition. To imitate this, we selected a set of diseases similar to the target disease; this was done by looking at other ICD10CM with the same parent code. When computing the similarity between symptom embedding and disease embedding composition, we used the phecode embedding of the alternative disease. Finally, we wanted to assess the situation where incorrect information is included in the symptom list. We simulated this by adding a randomly selected ICD10CM code to the symptom list and included it in the computation of the aggregated symptoms. Because we use vector addition to aggregate symptoms and addition is commutative, we place the random symptom at the beginning of the aggregation procedure.

### Phenotype risk score

#### Case/control construction

To define cases, we first selected all patients with at least one ICD9CM code from the list of inclusion codes (CHS encodes their EHR using ICD9CM). We then filter patients to include only patients listed on the CHS chronic registry to confirm they have received the disease diagnosis. Because patients are not guaranteed to be added to the chronic registry when they are first diagnosed, we treat the first occurrence of the inclusion ICD9CM code as the date of diagnosis. For each patient, we set the index date as January 1 of the year of diagnosis, where only information occurring before the index date will be included in the risk score computation. For prediction, we select all EHRs recorded within the past 5 years of the index date.

To define controls, we begin with the phecode exclusion criteria provided by the phecode mapping v1.2. For each phecode, the mapping provides a list of exclusion phecodes that are typically similar conditions or overlap with the target disease. We begin by selecting all individuals who have never received any exclusion diagnoses at any point in their EHR. We also remove individuals from the chronic disease registry for the specified disease. We select a matched control from this set of candidate control patients for each case. Matching is based on the following criteria: 1) same sex, 2) year of birth ±2 years, and 3) number of recorded diagnoses in the EHR (with at least 1 diagnosis in the EHR). The complete case-control process is illustrated in Supplementary Figure S5.

#### Risk score computation

For a given disease, a risk score is computed as the weighted sum of relevant clinical codes, where the weight is the cosine similarity between the embedding of each code and the disease embedding. We restricted the set of potential features to laboratory tests, medications, and ICD9CM codes with at least a 0.1% prevalence within the CHS patient population. The CHS EHR did not have SNOMED CT or CPT codes for analysis. To determine the clinical codes related to each disease, we compute the cosine similarity between all clinical codes within the latent space and the target disease embedding, represented by a phecode. The final set of features is the top k codes, where *k*=20, and the feature weights are the cosine similarity scores. In practice, the choice of *k* can be treated as a hyper-parameter and optimized with a validation dataset, similar to the p-value thresholding of polygenic risk scores. Patient risk scores are computed as a weighted sum where the associated weight is added to the sum if the patient has the given code in their EHR and given a 0 otherwise. Features are counted once in the score computation, even if a patient receives the codes multiple times.

#### Survival analysis

For each disease, we computed the most common age at diagnosis and the sex with the highest disease prevalence. We restricted the survival analysis to patients meeting these age/sex criteria and those with a diagnosis date from 2016 or earlier. We re-computed risk score quantiles based only on this restricted set of patients. We looked at the 6 years of EHR after each patient’s index date and denoted a patient as deceased if they had a non-null death date. Patients were stratified into risk score quartiles based on the risk score computed at the index date. Confidence intervals for the survival curves were computed using 10,000 bootstrapped samples.

### Integrating *ClinVec* with LLMs

We augment MMed-Llama ^106^—an 8-billion-parameter open-source medical foundation model that already incorporates domain-specific knowledge through pre-training—with 42,883 *ClinVec* embeddings and a 4-layer MLP projector module to align embedding dimensions which is trained with parameter efficient fine-tuning. We download the pre-trained model from Hugging

Face (https://huggingface.co/Henrychur/MMed-Llama-3-8B) and use the authors’ fine-tuning pipeline to perform parameter efficient fine-tuning with LoRA^140^ (https://github.com/MAGIC-AI4Med/MMedLM/tree/main). We used the same prompts and sets of hyper-parameters pro vided by MMed-Llama, specifically tuning the following target modules: ‘q_proj’, ‘v_proj’, ‘v_proj’, ‘mlp’. Fine-tuning was performed on the training English subset of multiple-choice questions from the MedQA dataset (https://huggingface.co/datasets/VodLM/medqa).

#### MetaMap

We annotate each multiple-choice question using the MetaMap^141^ tool, which annotates natural language text with UMLS CUI codes. For each question, we concatenated the text from the ques tion and the answer options and submitted the text to the MetaMapLite REST API (https://ii.nlm.nih.gov/metamaplite/rest). We limit the choice of UMLS CUI codes to the follow ing semantic types: “Sign or Symptom”, “Disease or Syndrome”, “Pharmacological Substance”, “Mental or Behavioral Dysfunction”, “Chemical”, “Anatomical Structure”.

#### AfriMed-QA

We also perform a validation study on the AfriMed-QA^109^ dataset (https://huggingface.co/datasets/intronhealth/afrimedqa_v2). We restricted to only multiple-choice questions and removed questions that had more than one assigned correct answer. The original dataset provides clinical subspecialty information for each question.

### Institutional Review Board approval

Parts of this study that relate to the use of CHS data (for evaluating the predictive ability of the KG-derived concept embeddings) were approved by the CHS Institutional Review Board (Hel sinki) committee.

## Data availability

Data and clinical concept embeddings, as well as the *ClinGraph* knowledge graph, are available via Harvard Dataverse https://doi.org/10.7910/DVN/Z6H1A8. Due to national and organiza tional data privacy regulations, CHS individual-level data from this study cannot be shared publicly.

## Code availability

The Python implementation of the heterogeneous graph transformer and the associated codebase for model training, (non-individual-level) analyses, and embedding evaluations are available at https://github.com/mims-harvard/Clinical-knowledge-embeddings. The project website is at https://zitniklab.hms.harvard.edu/projects/Clinical-knowledge-embeddings.

## Acknowledgements

R.J., B.Y.R., R.D.B., N.D., and M.Z. are supported by the Berkowitz Family Living Laboratory at Harvard Medical School and the Clalit Research Institute. R.J. and M.Z. gratefully acknowledge the support of NIH R01-HD108794, NSF CAREER 2339524, US DoD FA8702-15-D-0001, awards from Harvard Data Science Initiative, Amazon Faculty Research, Google Re search Scholar Program, AstraZeneca Research, Roche Alliance with Distinguished Scientists, Sanofi iDEA-iTECH, Pfizer Research, Chan Zuckerberg Initiative, John and Virginia Kaneb Fel lowship at Harvard Medical School, Biswas Computational Biology Initiative in partnership with the Milken Institute, Harvard Medical School Dean’s Innovation Fund for the Use of Artificial Intelligence, and Kempner Institute for the Study of Natural and Artificial Intelligence at Har vard University. Any opinions, findings, conclusions or recommendations expressed in this ma terial are those of the authors and do not necessarily reflect the views of the funders. We grate fully acknowledge the resources provided by the Clalit Research Institute. We thank Michelle M. Li, Ayush Noori, Intae Moon, and Shvat Messica for contributing to the model development and analyses.

## Author contributions

R.J. developed a unified knowledge graph representation of clinical concepts and designed, im plemented, and benchmarked heterogeneous graph transformer models. U.G. implemented the approach and conducted detailed analyses using CHS individual-level data. L.H., J.W., S.D., C.R.G., P.H., and R.S. formed an inter-institutional panel of clinicians that evaluated the align ment of clinical concept representations with established clinical knowledge across 90 diseases and 3,000 clinical codes. B.Y.R. and R.B. provided feedback on the methodology and evaluated the clinical concept embeddings through large-scale phenotype risk scoring and alignment with expert clinical knowledge. R.J., N.D., and M.Z. conceptualized and designed the study. All au thors contributed to writing the manuscript.

## Competing interests

The authors declare no competing interests.

## Notes

### Competing Interest Statement

The authors have declared no competing interest.

### Author Declarations

Parts of this study that relate to the use of Clalit Healthcare Services (CHS) data (for evaluating the predictive ability of the KG-derived concept embeddings) were approved by the CHS Institutional Review Board (Helsinki) committee.

### Summary of Updates

Updated to address reviewer comments.

