## Supplementary Materials for "ClinVec: Unified Embeddings of Clinical Codes Enable Knowledge-Grounded AI in Medicine"

**Figures:**


- **Supplementary Figure S1: Clustering of biomedical knowledge graph embeddings.**
- **Supplementary Figure S2: Embedding composition of autoimmune disorders.**
- **Supplementary Figure S3: Embedding composition of leading non-communicable diseases.**
- **Supplementary Figure S4: Perturbations of embedding composition of leading non-communicable diseases.**
- **Supplementary Figure S5: Flowchart outlining case and control cohort construction for phenotype risk scores.**
- **Supplementary Figure S6: Clinical code embedding risk scores correlate with disease severity.**
- **Supplementary Figure S7: Overview of clinical knowledge embedding integration for medical question answering.**
- **Supplementary Figure S8: Improved medical question answering performance with clinical embeddings.**


**Tables:**


- **Supplementary Table S1: Summary of node types in *ClinGraph*.**
- **Supplementary Table S2: Summary of *ClinGraph* edges**.
- **Supplementary Table S3: Comparison of EHR vocabularies across biomedical knowledge graphs.**
- **Supplementary Table S4A: Zero-shot disease-symptom retrieval across embedding frameworks**
- **Supplementary Table S4B: Zero-shot disease indication retrieval across embedding frameworks.**
- **Supplementary Table S5: Symptom lists and corresponding ICD-10 codes.**
- **Supplementary Table S6: Summary statistics of case and control cohorts for phenotype risk prediction.**
- **Supplementary Table S7: Cosine similarity between aggregated symptoms and disease embeddings.**

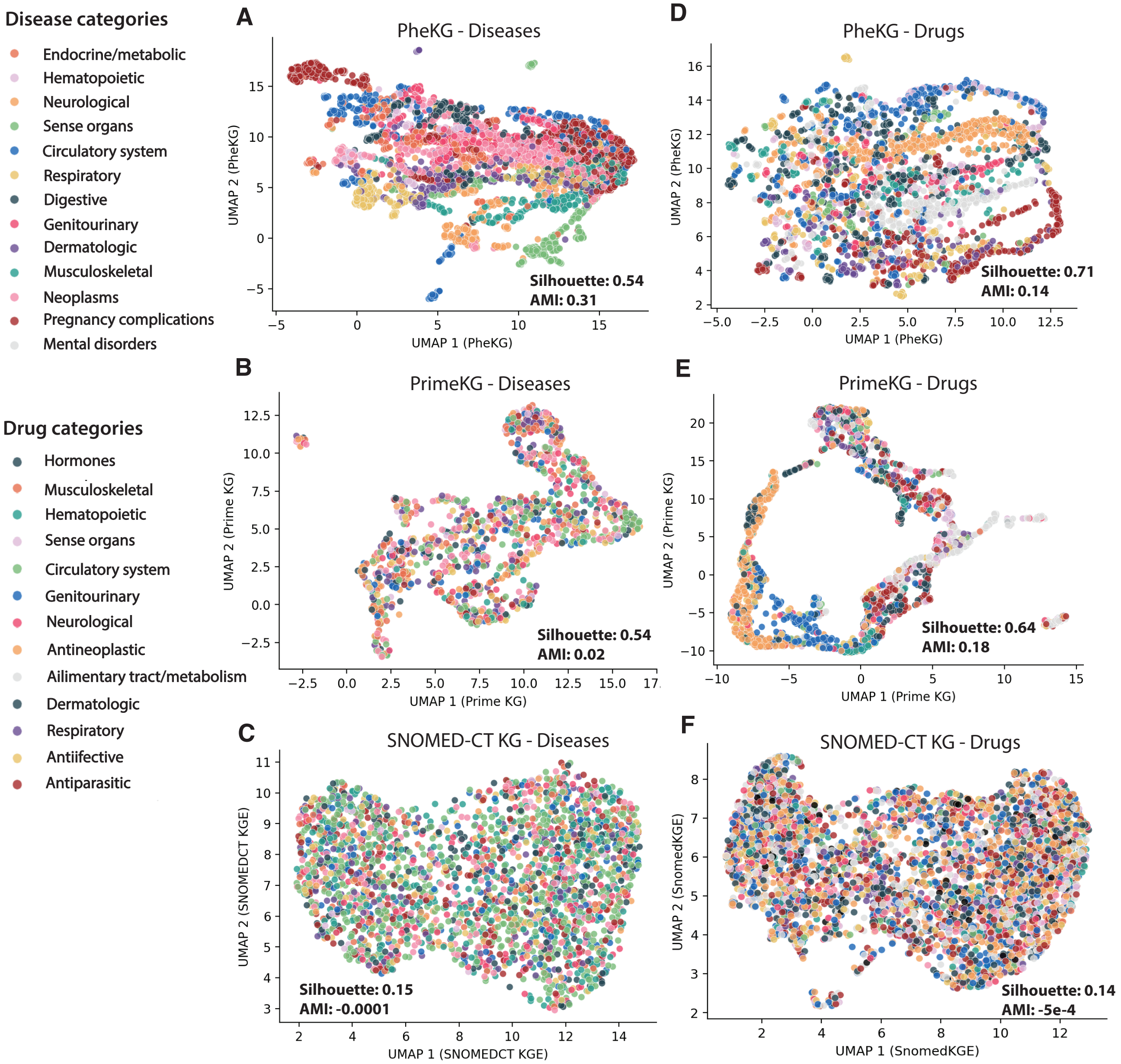


**Supplementary Figure S1: Clustering of biomedical knowledge graph embeddings.** We provide plots of the first two dimensions of the latent embedding spaces defined by embeddings produced by *ClinGraph* (*ClinVec*, first column), PrimeKG (middle), and SNOMED-CT KG (last). (A-C) show clustering by diagnosis code category and (D-F) show clustering by ATC-1 category. Markers are shaded according to their respective categories. For each set of embeddings, we report the silhouette score and the adjusted mutual information (AMI).

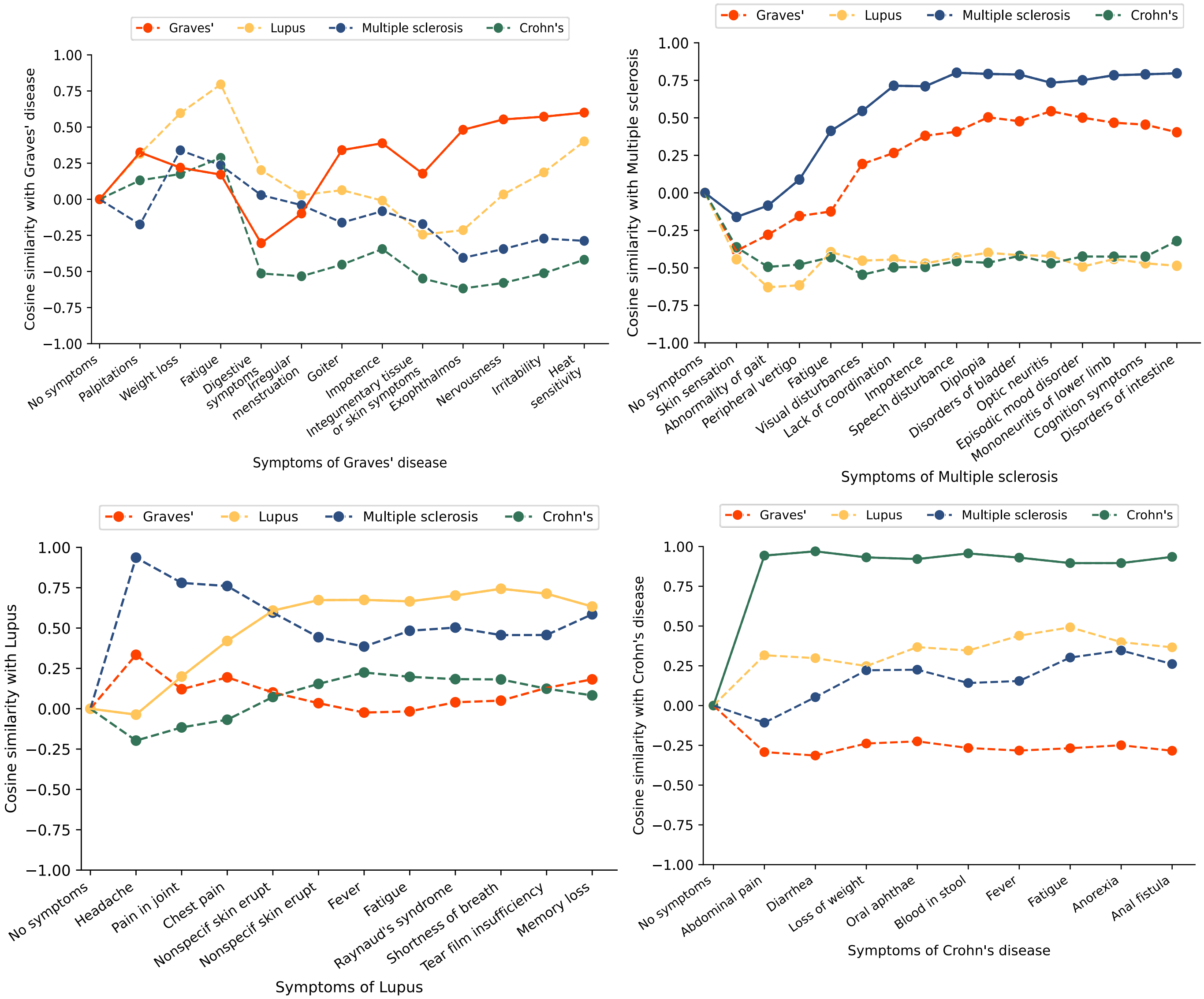


**Supplementary Figure S2: Embedding composition of autoimmune disorders.** Line plot showing the cosine similarity across 4 autoimmune diseases and the aggregation of Graves' disease (red), multiple sclerosis (blue), lupus (yellow), and Crohn's disease (green) symptoms. Aggregated embeddings are computed by sum pooling across the embeddings for each symptom and ordered on the x-axis according to population frequency within the Clalit Healthcare system.

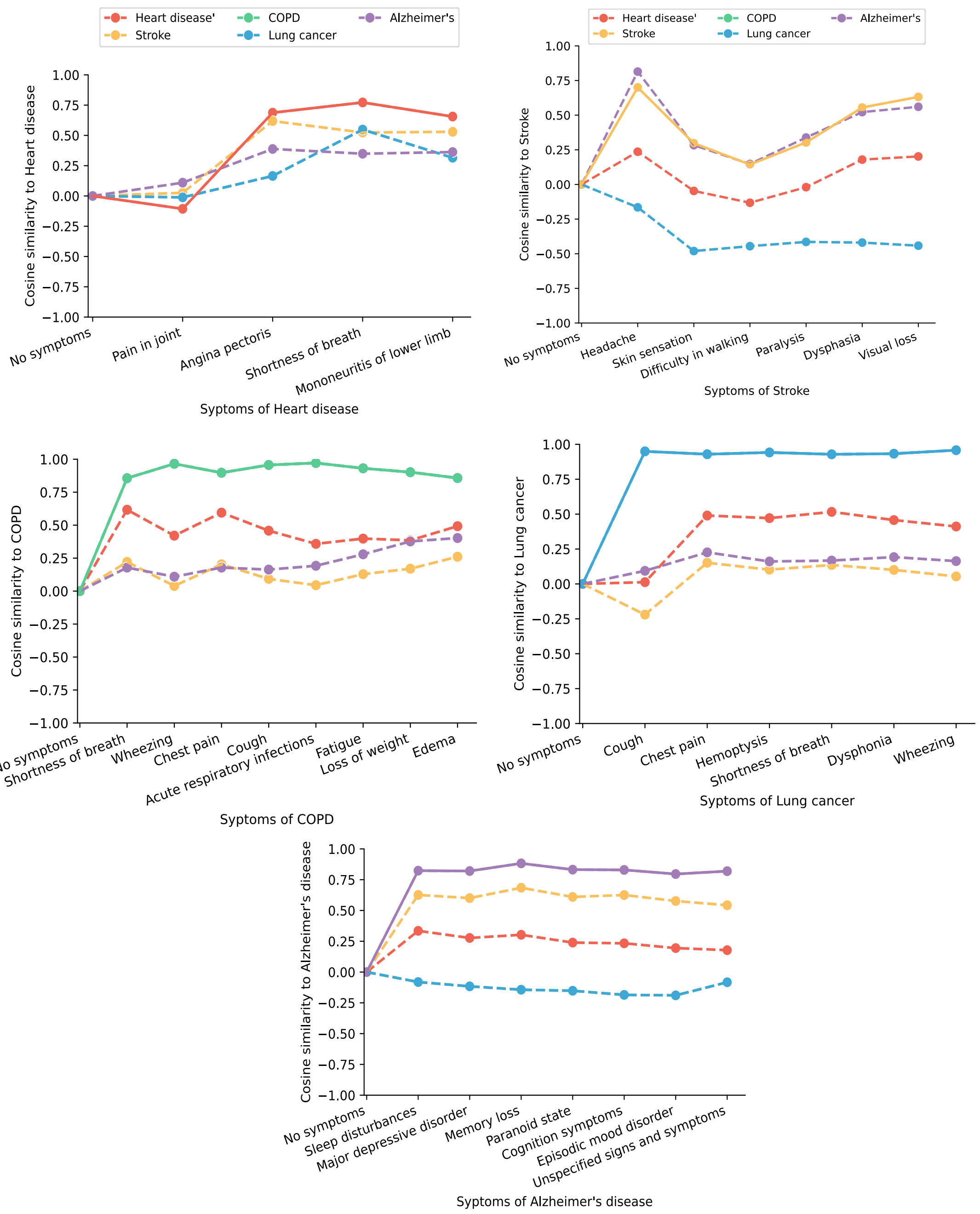


**Supplementary Figure S3: Embedding composition of leading non-communicable diseases.** Line plot showing the cosine similarity across 5 non-communicable diseases and the aggregation of heart disease (red), stroke (yellow), COPD (green), lung cancer (blue), and Alzheimer's disease (purple). Aggregated *ClinVec* embeddings are computed by sum pooling across the embeddings for each symptom and ordered on the x-axis according to population frequency within the Clalit Healthcare system.

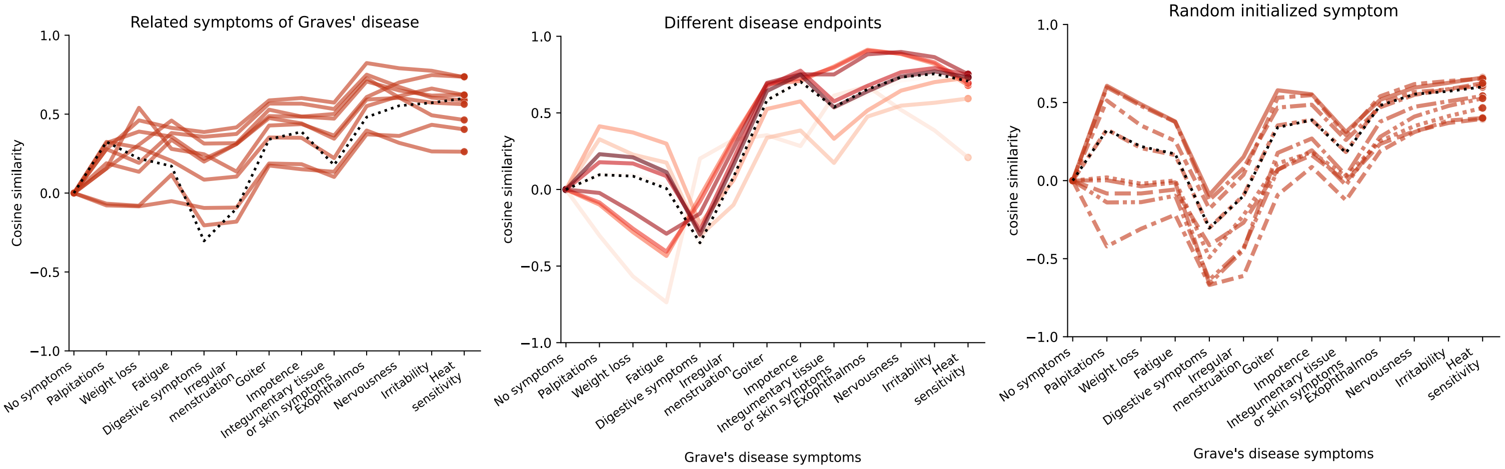


**Supplementary Figure S4: Perturbations of embedding composition of leading non-communicable diseases.** Line plot showing the cosine similarity of Graves' disease versus the aggregation of Graves' disease symptoms with varying levels of perturbation. Aggregated *ClinVec* embeddings are computed by sum pooling across the embeddings for each symptom and ordered on the x-axis according to population frequency within the Clalit Healthcare system. (A) At each symptom, we sample a related ICD-10 code in the computation of the composition embedding. (B) We use 10 diseases related to Graves' disease and use this as the endpoint for each disease embedding. (C) We initialized the disease composition embedding with a randomly selected ICD-9 code. Each process is repeated for 10 times, yielding 10 curves for each setup.

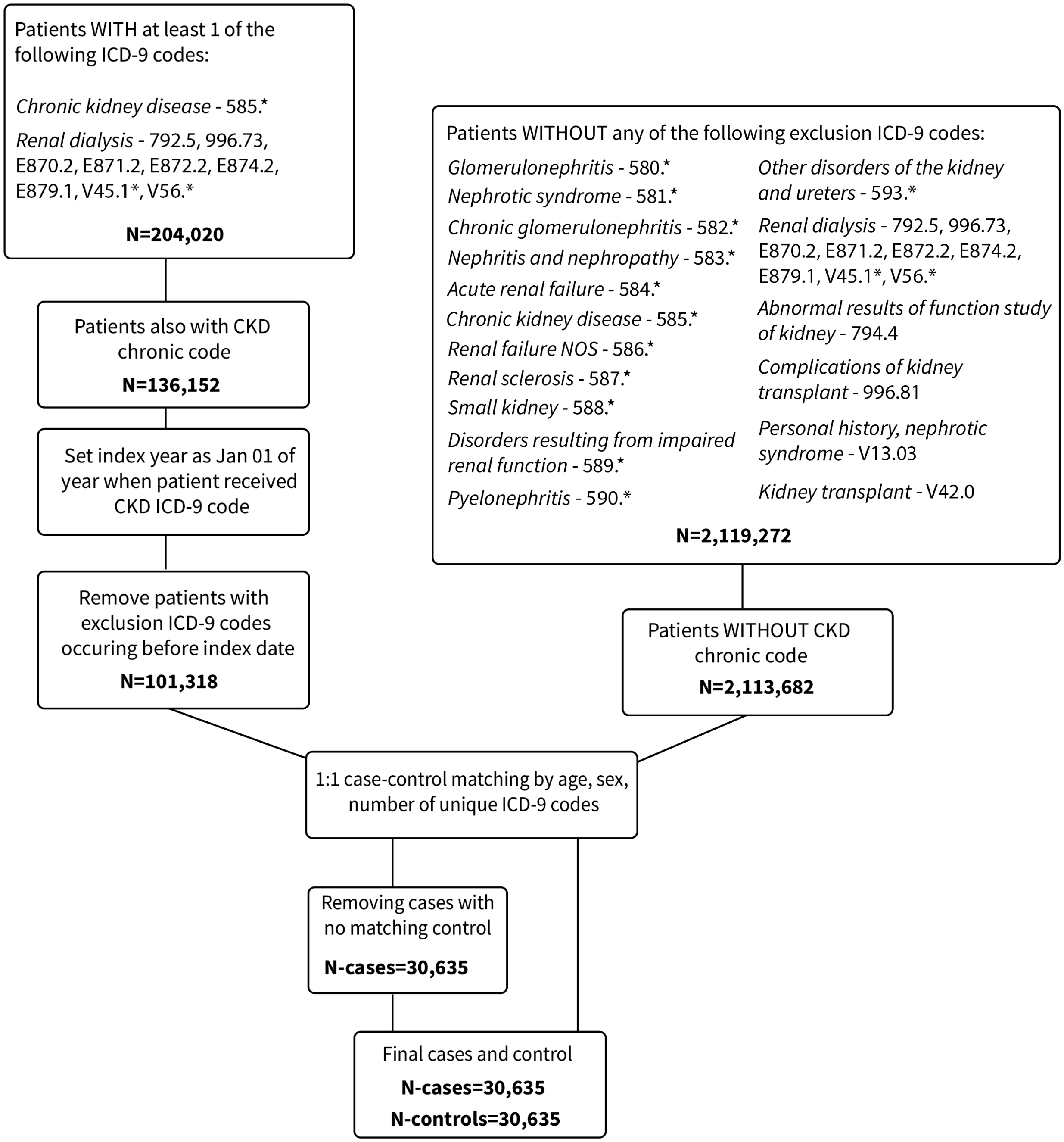


**Supplementary Figure S5: Flowchart outlining case and control cohort construction for phenotype risk scores.** We outline the steps used to construct case-control cohorts for chronic kidney disease as an example. 'Chronic codes' refer to codes placed on patient's records indicating membership in CHS's chronic disease registry. The set of exclusion codes is determined by the provided exclusion list featured in the phecode v1.2 definitions.

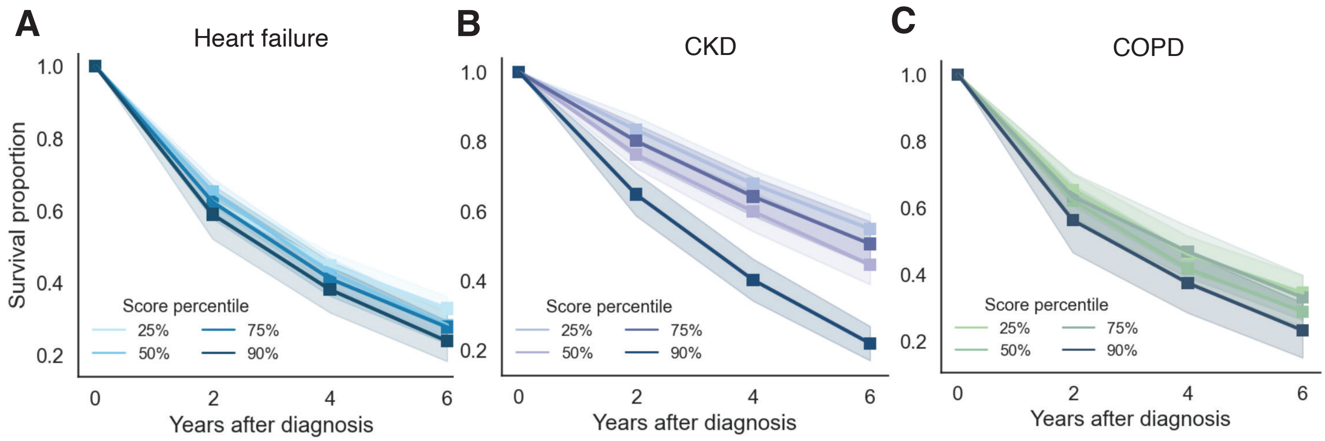


**Supplementary Figure S6: Clinical code embedding risk scores correlate with disease severity.** We show survival curve of cases that match the most common sex and age at diagnosis for each disease cohort. Lines are shaded by score percentiles. Standard error intervals are computed using 10,000 bootstrap samples. Columns are divided by diseases: heart failure, chronic kidney disease, and COPD.

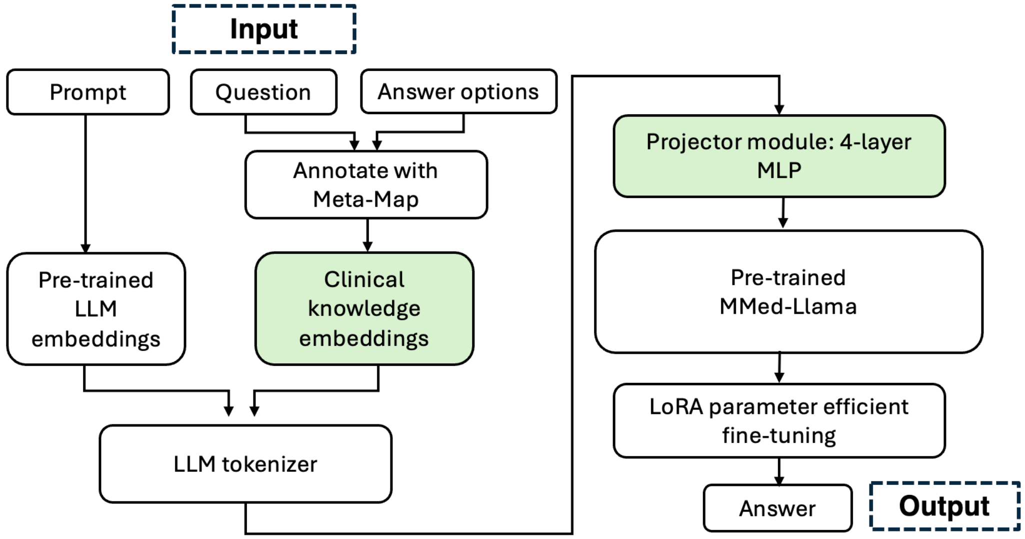


**Supplementary Figure S7: Overview of clinical knowledge embedding integration for medical question answering.** We show a flowchart summarizing how *ClinVec* embeddings are integrated with pre-trained LLMs. Boxes in green denote the specific steps involving clinical knowledge integration.

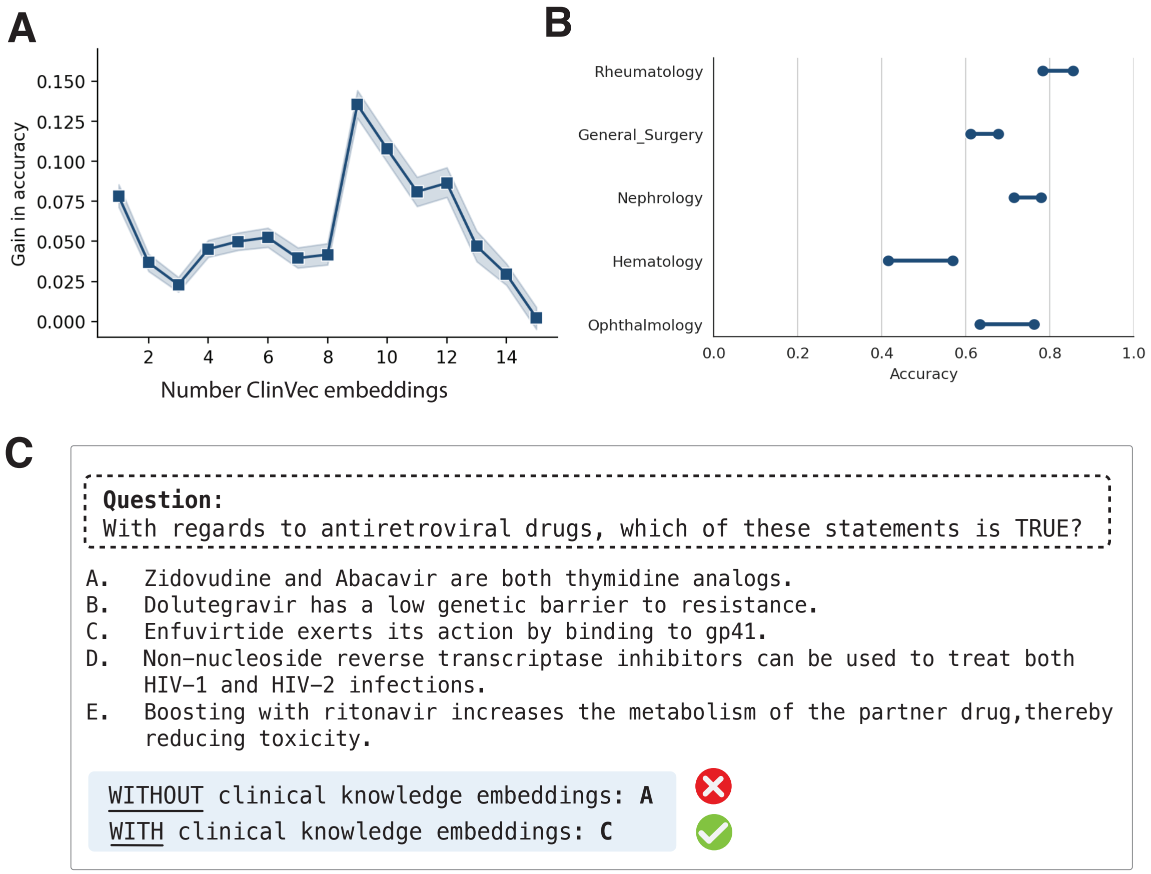


**Supplementary Figure S8: Improved medical question answering performance with clinical embeddings.** (A) Questions are binned according to the number included *ClinVec* embeddings (as assigned by MetaMap). (B) Accuracy of N=3,724 multiple-choice questions from AfriMed-QA. The red line denotes random performance with 5 answer options. (C) Stratified performance across questions of different clinical specialties. The right- and left-hand markers denote performance with and without the inclusion of *ClinVec* and the connecting line denotes the difference in absolute performance. Standard errors are estimated from bootstrapped samples. (D) Sample hematology question from the AfriMed-QA dataset answered correctly when including *ClinVec*.

| Node type | Description | Number of nodes |
|---|---|---|
| ATC | Anatomical Therapeutic Chemical (ATC) Classification codes provide a hierarchical classification for pharmaceutical products. | 5,179 |
| CPT | Current Procedural Terminology (CPT®) codes represent a unifying nomenclature describing medical, surgical, and diagnostic services. | 4,377 |
| ICD9CM | The International Classification of Diseases 9th Revision (ICD-9-CM) system is a unifying coding system for medical diagnoses. It is also the main medical coding system used for billing and claims reimbursement. | 10,157 |
| ICD10CM | The International Classification of Diseases 10th Revision (ICD-10-CM). We restrict inclusion to codes with <=5 digits. | 34,842 |
| LOINC | Logical Observation Identifiers Names and Codes (LOINC) is a coding database that provides a universal set of names and codes for laboratory and clinical test results. LOINC Parts are coded representations of individual terms that make up a LOINC term. | 14,399 |
| Phecode | Phecodes are an aggregation schema for ICD-9-CM codes that represent clinically meaningful distinct diseases and traits for primary clinical or genetic research. Here we use v1.2. | 1,793 |
| RxNorm | RxNorm provides normalized names for generic and branded clinical drugs. | 10,130 |
| SNOMED CT | SNOMED CT US & SNOMED Clinical Terms (CT) is a standardized set of medical terms providing codes, terms, synonyms and definitions used in clinical documentation and reporting. | 29,406 |
| UMLS CUI | The Unified Medical Language System (UMLS) is organized by the National Library of Medicine and integrates multiple clinical vocabularies and coding systems into a single terminology. Each concept is assigned a concept unique identifier (CUI) code. | 42,883 |

**Supplementary Table S1: Summary of node types in *ClinGraph*.** We provide the description and the number of nodes per clinical vocabulary in *ClinGraph*.

| **Edges** | **N** | **Knowledge sources** |
|---|---|---|
| *UMLS-CUI <> ICD10CM* | 5,066 | UMLS |
| *UMLS-CUI <> ICD9CM* | 2,992 | |
| *UMLS-CUI <> SNOMEDCT* | 15,746 | |
| *UMLS-CUI <> RXNORM* | 2,532 | |
| *UMLS-CUI <> ATC* | 2,670 | |
| *UMLS-CUI <> LNC* | 7,288 | |
| *UMLS-CUI <> CPT* | 188 | |
| *PHECODE <> ICD10CM* | 1,565,558 | Phecodes v1.2, PheMap |
| *PHECODE <> ICD9CM* | 488,298 | |
| *UMLS-CUI <> PHECODE* | 281,230 | PheMap |
| *PHECODE <> SNOMEDCT* | 62,372 | |
| *PHECODE <> CPT* | 4,878 | |
| *PHECODE <> LNC* | 48,302 | |
| *PHECODE <> RXNORM* | 7,334 | |
| *ATC<>RXNORM* | 37,140 | ATCProd |
| *ICD9CM <> ICD10CM* | 29,552 | GEM |
| *SNOMEDCT <> ICD10CM* | 112,000 | SNOMEDCT US |
| *LNC<>LNC* | 29,652 | UMLS |
| *CPT<>CPT* | 11,260 | |
| *ATC<>ATC* | 8,758 | |
| *ATC <> ICD10CM* | 8,274 | |
| *LNC<>ICD10CM* | 3,988 | |

**Supplementary Table S2: Summary of *ClinGraph* edges**. We provide an overview of the top number and type of edges present in *ClinGraph*. In the far-right column, we list the source databases for the relations.

| | *Diseases % (ICD10)* | *Diseases % (ICD10-root)* | *Drugs % (ATC5)* | *LOINC %* | *CPT %* | *SNOMEDCT US %* |
|---|---|---|---|---|---|---|
| PrimeKG[4] | 23.12* | 57.23* | 52.55* | 0 | 0 | 0 |
| DRKG[13] | 1.34* | 52.51* | 39.31* | 0 | 0 | 0 |
| PharMeBINet[14] | 22.48* | 54.81* | 33.78* | 0 | 0 | 0 |
| Hetionet[15] | 0.09* | 10.98* | 24.20* | 0 | 0 | 0 |
| GAT-ETM[16] | 100 | 100 | 100 | 0 | 0 | 0 |
| SNOMED-KGE[3] | 0 | 0 | 0 | 0 | 0 | 47.17 |
| **ClinVec** | 25.17 | 85.86 | 81.28 | 20.57 | 39.17 | 6.28 |

**Supplementary Table S3: Comparison of EHR vocabularies across biomedical knowledge graphs.** We report the percentage of vocabulary covered by each knowledge graph (KG). KGs that are not originally represented by EHR vocabularies are mapped using pre-defined mappings (see Methods). These statistics are marked with a (*).

### Hits @ K - Disease symptoms

| | *K=3* | *K=5* | *K=25* | *K=50* | *K=100* |
|---|---|---|---|---|---|
| Snomed2vec | 0 | 0 | 0.50 | 0.667 | 0.833 |
| Cui2vec | 0.333 | 0.333 | 0.833 | 0.833 | 0.833 |
| **ClinVec** | 0.50 | 0.667 | 0.833 | 0.833 | 1.0 |

**Supplementary Table S4A: Zero-shot disease-symptom retrieval across embedding frameworks.** For each embedding method, we report the hits @ k computed over 6 diseases. We define hits @ k as the proportion of diseases for which a relevant symptom appears within the top k retrieved codes.

### Hits @ K - Drug indications

| | *K=3* | *K=5* | *K=25* | *K=50* | *K=100* |
|---|---|---|---|---|---|
| Snomed2vec | 0 | 0 | 0 | 1.0 | 0.33 |
| Cui2vec | 0.167 | 0.50 | 0.667 | 0.833 | 0.833 |
| **ClinVec** | 0.167 | 0.50 | 0.833 | 1.0 | 1.0 |

**Supplementary Table S4B: Zero-shot disease indication retrieval across embedding frameworks.** For each embedding method, we report the hits @ k computed over 6 diseases. We define hits @ k as the proportion of diseases for which a relevant medication code appears within the top k retrieved codes.

[see Excel sheet]

**Supplementary Table S5: Symptom lists and corresponding ICD-10 codes.** For a given disease, we retrieve the symptom list listed on the Mayo Clinic disease database. Each symptom is then translated by hand to a corresponding ICD-10 code. The diseases include: Graves' disease, lupus, multiple sclerosis, and Crohn's disease, heart disease, lung cancer, Alzheimer's disease, and chronic obstructive pulmonary disease (COPD).

| **Disease** | | **N** | **Mean age (SD)** | **Sex** | **ICD-9 codes (EHR length)** |
| --- | --- | --- | --- | --- | --- |
| Heart failure | *Case* | 20,452 | 76.2 (11.5) | M: 50.2%<br>F: 49.8% | 300.6 (195.7) |
| | *Control* | 26,266 | 76.2 (11.8) | M: 51.9%<br>F: 48.1% | 273.3 (188.8) |
| Chronic obstructive pulmonary disease (COPD) | *Case* | 2,176 | 71.0 (12.3) | M: 64.6%<br>F: 35.4% | 278.7 (223.4) |
| | *Control* | 9,438 | 69.1 (12.8) | M: 68.2%<br>F: 31.8% | 225.6 (198.1) |
| Chronic kidney disease (CKD) | *Case* | 10,369 | 74.1 (11.4) | M: 56.3%<br>F: 43.7% | 271.7 (190.7) |
| | *Control* | 12,802 | 74.2 (11.7) | M: 57.3%<br>F: 42.7% | 237.4 (174.7) |

**Supplementary Table S6: Summary statistics of case and control cohorts for phenotype risk prediction.** For each disease, across cases and controls, we list the sample size, mean age, standard deviation of age, sex, and length of EHR. The EHR-length is computed as the total number of unique ICD-9 codes on a patient's record prior to the index date.

| | | Disease embedding | | | | | | | | |
|---|---|---|---|---|---|---|---|---|---|---|
| | | *COPD* | *Crohn's* | *Dementia* | *Graves'* | *Heart disease* | *Lung cancer* | *Lupus* | *MS* | *Stroke* |
| **Aggregated symptoms** | *COPD* | -0.016 | 0.646 | 0.257 | 0.397 | -0.538 | 0.600 | 0.713 | 0.695 | 0.289 |
| | *Crohn's* | 0.154 | 0.908 | 0.181 | 0.453 | -0.243 | 0.643 | 0.560 | 0.575 | -0.035 |
| | *Dementia* | -0.265 | 0.126 | 0.909 | 0.727 | -0.233 | -0.266 | -0.032 | 0.330 | 0.443 |
| | *Graves'* | -0.197 | 0.632 | 0.616 | 0.765 | -0.402 | 0.242 | 0.477 | 0.684 | 0.328 |
| | *Heart disease* | 0.326 | 0.481 | -0.263 | -0.249 | 0.583 | 0.096 | -0.280 | -0.388 | -0.324 |
| | *Lung cancer* | 0.237 | 0.638 | -0.012 | 0.164 | -0.370 | 0.759 | 0.631 | 0.508 | 0.045 |
| | *Lupus* | -0.316 | 0.467 | 0.064 | 0.515 | -0.723 | 0.544 | 0.988 | 0.930 | 0.377 |
| | *MS* | -0.543 | 0.298 | 0.803 | 0.816 | -0.602 | -0.114 | 0.440 | 0.744 | 0.560 |
| | *Stroke* | -0.732 | -0.267 | 0.888 | 0.694 | -0.458 | -0.651 | 0.028 | 0.421 | 0.650 |

**Supplementary Table S7: Cosine similarity between aggregated symptoms and disease embeddings.** For each disease, we compute the cosine similarity between a disease embedding (columns) and the aggregated symptom embeddings (rows). Symptoms are aggregated using sum-pooling.

| UMLS CUI | UMLS CUI name | Phecode | Phecode name | Symptoms |
|---|---|---|---|---|
| C0152105 | Hypertensive heart disease | 411.8 | Other chronic ischemic heart disease, unspecified | 'C1998435', 'C0002962', 'C0015672', 'C0235880', 'C0154747', 'C0013404', 'C4075947' |
| C0038454 | cerebrovascular accident | 433.21 | Occlusion and stenosis of vertebral artery w/ cerebral infarction | 'C0973461', 'C0003537', 'C4039303', 'C0030554', 'C0522224', 'C0427007', 'C2349426', 'C0018681', 'C4074978', 'C0311394' |
| C0024121 | Lung neoplasm | 165.1 | Cancer of bronchus; lung | 'C0742857', 'C0010200', 'C0008031', 'C0522051', 'C0043144', 'C0019079' |
| C0002395 | ALZHEIMER DISEASE | 290.11 | Alzheimer's disease | 'C0002622', 'C0751295', 'C0557932', 'C0154440', 'C0152125', 'C1456784', 'C0150079', 'C0037317', 'C0085633', 'C0231686' |
| C0010346 | CROHN DISEASE | 555.1 | Regional enteritis | 'C0474496', 'C0239181', 'C0235880', 'C0154747', 'C0426587', 'C0232462', 'C4047940', 'C0018932', 'C1321898' |
| C0026769 | Sclerosis multiple | 335.0 | Multiple sclerosis | 'C2316899', 'C0575081', 'C0235880', 'C0154747', 'C0239652', 'C0030554', 'C0861152', 'C0029134', 'C0015672', 'C3661940' |

| UMLS CUI | UMLS CUI name | Phecode | Phecode name | Drugs |
|---|---|---|---|---|
| C0024305 | lymphoma, non-Hodgkin's | 202.2 | Non-Hodgkins lymphoma | 'C0010583', 'C0013089', 'C0042679', 'C0032952' |
| C0006413 | Burkitt lymphoma | n/a | n/a | 'L01AA01', 'L01DB01', 'L01CA02', 'A07EA03', 'H02AB07' |
| C0153594 | Malignant neoplasm of testis | 187.2 | Malignant neoplasm of testis | 'C0005740', 'L01DC01', 'C0015133' |
| | | | | 'L01CB01', 'C0008838', 'L01XA01' |
| C1269683 | MAJOR DEPRESSIVE DISORDER | 296.22 | Major depressive disorder | 'C0008845', 'C0074393', 'C1099456', 'C0016365', 'C0070122' |
| | | | | 'N06AB04', 'N06AB06', 'N06AB10', 'N06AB03', 'N06AB05' |
| C0005586 | Bipolar affective disorder | 296.1 | Bipolar | 'C0023870', 'C0080356', 'C0006949', 'C0064636', 'C0171023','C0018546', 'C0123091' |
| | | | | 'N05AN01', 'N03AG01', 'N03AF01', 'N03AX09', 'N05AH03', 'N05AD01', 'N05AH04' |

| | | | | |
|---|---|---|---|---|
| C0036341 | Schizophrenia | 295.1 | Schizophrenia | 'C0008286', 'C0016368', 'C0018546', 'C0031184' |
| | | | | 'N05AA01', 'N05AB02', 'N05AD01', 'N05AB03' |

**Supplementary Table S8: List of disease symptoms and drug indications for retrieval comparison.** We provide the list of diseases (both in UMLS CUI and phecode format) used in the retrieval @ k evaluation tasks. The first section lists the corresponding disease symptoms in UMLS CUI format. The second section lists the drugs used in each first-line treatment. In addition to drugs in UMLS CUI format, we also provide the corresponding ATC5 code in the row below.
